# An Agent-Based Modeling Framework for Healthcare AI Adoption: Application to Ambient Clinical Documentation

**DOI:** 10.64898/2026.07.01.26357077

**Authors:** Matthew G. Crowson

## Abstract

**Objective:** To develop and demonstrate an agent-based modeling framework for healthcare AI adoption, using ambient clinical documentation as the calibration case.

**Materials and Methods:** We built an agent-based model with 50,000 clinician agents, 500 organization agents, and 4 vendor agents over 104 weeks. Modeled clinicians differed by psychotype, specialty, and friction/benefit thresholds; modeled organizations progressed through deployment phases with governance delays anchored to 8-28 weeks. All cited deployment values were independently re-verified against primary sources, and the model was validated against published data from seven health systems and three benchmarks using formal goodness-of-fit metrics (RMSE, 90% predictive-interval coverage) grouped by reference class. After correcting an organization-initialization artifact, we performed a formal six-parameter re-calibration to align both the early-time trajectory and the steady-state plateau with published data. Six intervention scenarios were compared in paired simulations (n=30 realizations per scenario) using effect sizes with bootstrap intervals, and both the full intervention comparison and the Sobol sensitivity screen were re-run natively under the re-calibrated model. Global sensitivity analysis used Sobol indices (64 base samples; 1,152 parameter sets) across eight parameters.

**Results:** Baseline simulations produced S-curve adoption trajectories with wide variability. The re-calibrated model reproduced both early-time single-site trajectories and the cross-sectional adoption plateau, covering 86% of reference-class-matched anchors at the nominal 90% level, versus 29% for the original configuration. Most intervention scenarios increased adoption; in the original configuration the combined intervention outperformed individual levers, with significant interactions confirmed by 2^3^ factorial analysis. Re-running the analyses natively under the calibrated model both confirmed and revised these conclusions: governance remained the largest single structural lever and non-success absorbing states remained prominent, but intervention effects attenuated sharply, the combined intervention no longer reliably exceeded the best single lever at operating scale, and the leading sensitivity driver shifted from governance delay to clinician friction/edit-rate tolerance. That calibration changes which levers appear influential is itself the central methodological finding. Organizational outcomes clustered into non-success absorbing states (pilot stagnation and failure) alongside success and scaling.

**Conclusions:** Governance delay is an explicit upstream gate in the model, so its influence reflects model architecture and should not be interpreted as a universal real-world priority. The modeled pilot stagnation state is hypothesis-generating rather than an empirical category. Agent-based modeling provides a structured framework for understanding healthcare AI adoption dynamics. The approach supports hypothesis generation and comparative scenario exploration rather than point prediction.

## Introduction

Artificial intelligence is transitioning from research promise to clinical reality. Ambient documentation systems – AI tools that automatically generate clinical notes from patient-clinician conversations – represent one of the first widely deployed AI technologies in routine clinical practice. A 2024 survey of 43 health systems found ambient documentation to be the only AI use case with universal adoption activity [1], and major health systems have launched enterprise deployments with thousands of physicians [2], with adoption occurring faster than AI adoption in the broader economy [3]. Yet healthcare AI adoption remains poorly understood.

Despite substantial investment, early evidence suggests significant heterogeneity in outcomes: clinician-level uptake ranges from 20% to 74% across published implementations [4,5], with substantial pilot-to-scale attrition including 41% combined discontinuation and low utilization in one cohort study [5]. Industry assessments suggest many healthcare AI projects remain in early testing or pilot phases [6], which we model as a pilot stagnation state to represent stalled pilots. Traditional diffusion models, developed for consumer products, do not capture the complexity of healthcare settings where adoption involves individual clinician preferences, peer network effects (empirically estimated at 5.9-8.3% adoption increase per 10-point peer adoption increase [7]), organizational governance, and technical integration. There are limited quantitative frameworks for assessing which interventions may most effectively accelerate adoption and whether their effects interact.

We present an agent-based modeling framework for healthcare AI adoption, demonstrated through application to ambient documentation systems. Agent-based models capture micro-level adopter heterogeneity, meso-level peer influence dynamics, and macro-level organizational outcomes that emerge from their interaction [8]. The framework enables systematic comparison of intervention strategies, global sensitivity analysis, and characterization of emergent phenomena. While applied here to ambient scribes as a calibration case, the framework is intended to generalize to other healthcare AI technologies.

## Methods

### Model Overview

We developed an agent-based model following the ODD protocol [9], simulating weekly interactions among clinicians, organizations, and vendors over 104 weeks (**Figure 1**). The complete ODD protocol, parameter tables, and sensitivity analysis methodology are in Supplementary Methods.

**Figure 1.**
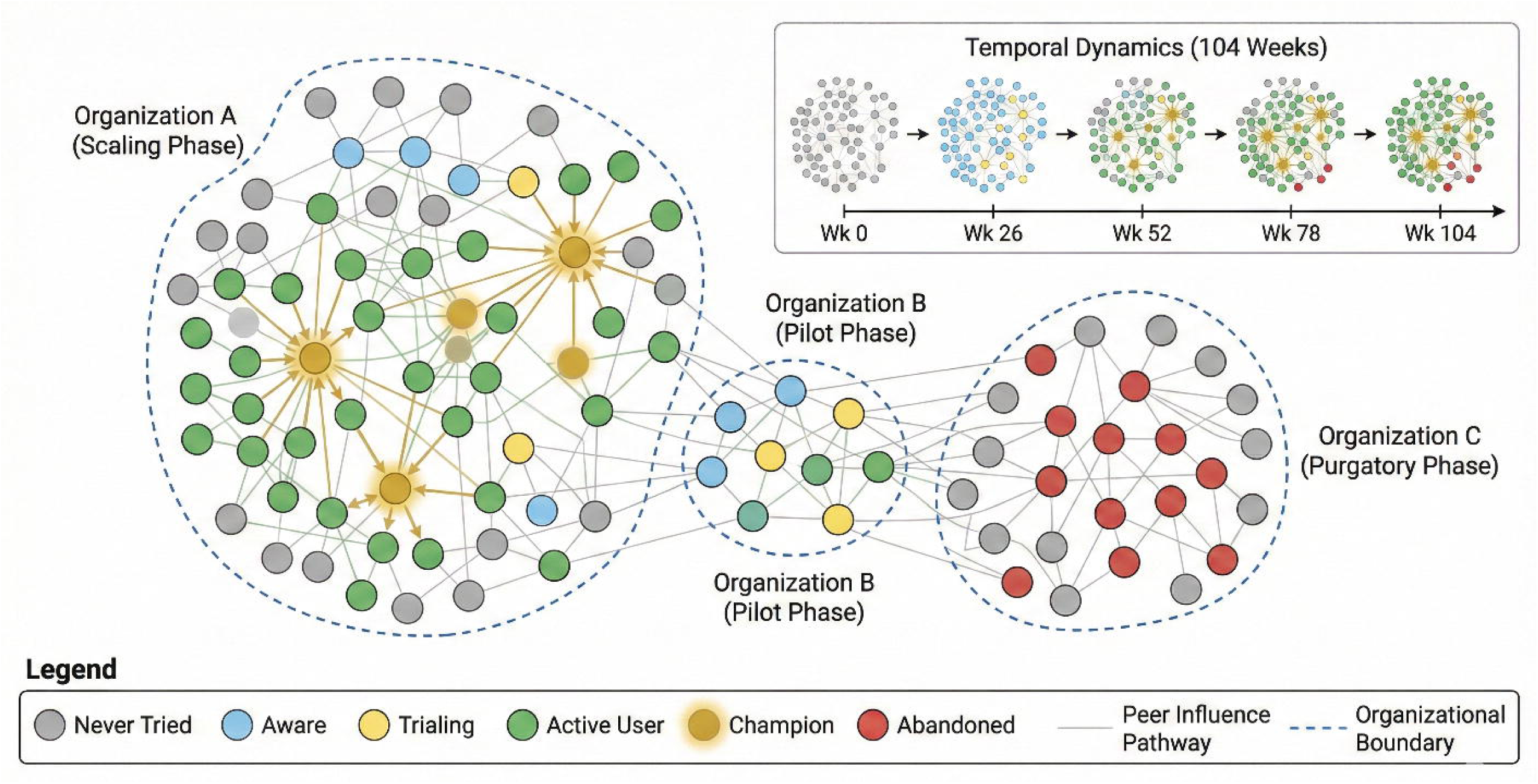
Model Overview Schematic representation of the agent-based model structure showing organizational deployment phases and clinician adoption states. Three representative organizations illustrate different deployment archetypes at week 104: Organization A (Scaling Phase) with high champion density and active peer influence cascades; Organization B (Pilot Phase) with early adoption activity and emerging trials; Organization C (Stagnation Phase) with stalled adoption and elevated abandonment. Node colors indicate clinician states: gray (never tried), blue (aware), yellow (trialing), green (active user), gold with rays (champion), red (abandoned). Edges represent peer influence pathways within the small-world network structure. Inset shows temporal dynamics of adoption spread across the 104-week simulation horizon.

### Agent Types and State Variables

**Clinician agents** (n=50,000) are characterized by: (1) an adoption state (never_tried, aware_not_tried, trialing, high_user, medium_user, low_user, champion, abandoned, or detractor); (2) a psychotype adapted from Rogers’ diffusion categories [10] (high-need early adopter 15%, evidence-driven pragmatist 40%, compliance-oriented late adopter 30%, resistant non-adopter 15%); (3) clinical specialty (primary care 40%, specialty medical 30%, surgical 20%, emergency 10%); and (4) individual thresholds for friction tolerance and minimum perceived benefit. Active use is defined as high_user, medium_user, low_user, or champion; adoption rate is the fraction of clinicians in active use states.

**Organization agents** (n=500) govern clinician access through deployment phases: not_started, evaluating, security_review, governance, pilot, scaling, and full_deployment (plus shadow_it for informal early use), with possible transitions to pilot stagnation or termination. Organizations vary in security review duration (12-26 weeks), governance delay (8-28 weeks, aligned to the ∼6.6-month procurement average [11]), integration maturity, and pilot success thresholds. The model supports two initialization regimes. In the default cold-start regime all organizations begin in a not-started state, producing near-zero ecosystem-level adoption in the first ∼10 weeks; this is a deliberately conservative assumption that, however, prevents comparison with published single-site trajectories observed mid-rollout. In an optional staggered regime, organizations are seeded at t=0 into a distribution of deployment phases (and, for organizations past governance, a phase-consistent distribution of clinician adoption states), calibrated to verified governance-to-deployment timelines from real systems (Cleveland ∼6 months, Vanderbilt ∼10 months, Kaiser ∼12 months, Mass General Brigham ∼24 months; Supplementary Methods S8). The staggered regime removes the cold-start artifact and enables formal comparison of early-time dynamics; both regimes are reported (Results; **Table 3**, **Figure 4**).

**Vendor agents** (n=4) represent ambient scribe products with varying performance, EHR integration quality, and pricing ($9,000-11,000/year [12]). Organizations select vendors through weighted evaluation; clinician experience is shaped by the organization-selected vendor’s attributes.

### Decision Rules

Clinician adoption decisions follow a utility-based framework. Key inputs include peer influence (5.9–8.3% per 10-point peer adoption increase [7]), experience accumulation over ∼10 trial encounters, edit-rate tolerance calibrated to ∼40% [13], and learning curves consistent with hybrid ambient documentation programs [14]. Champions emerge after 12 weeks of sustained positive experience and exert a 2× influence multiplier. Each clinician’s edit-rate tolerance is drawn from Normal(0.45, 0.08); exceeding tolerance triggers abandonment during trials and contributes to friction-driven reconsideration post-adoption. Detailed specifications for burnout dynamics, habit formation, and pilot enrollment are provided in Supplementary Methods S11.

Organization decisions follow sequential gating: evaluation → security review → governance → pilot. Pilot retention <0.20 triggers termination, 0.20–0.55 enters pilot stagnation, and ≥0.55 proceeds to scaling. Organizations in pilot stagnation exit via a retention- and time-dependent hazard. At simulation end, organizations are classified as success, scaling, pilot stagnation, stalled, failure, or unknown; thresholds are model-defined rather than empirically derived.

### Network Structure

Clinicians are embedded in a Watts-Strogatz small-world network (k=6, rewiring probability=0.1) with organization-based clustering overlay, consistent with the structure of physician professional networks [15]. The specific parameters produce a mean clustering coefficient of ∼0.4 and mean path length of ∼6.5 hops.

### Model Execution and Cross-Level Coupling

The simulation advances in weekly time steps; within each week the three agent levels are updated in a fixed order so that organizational state gates clinician behavior, and aggregated clinician behavior in turn feeds organizational decisions the following week (full scheduling in Supplementary Methods S1.3):

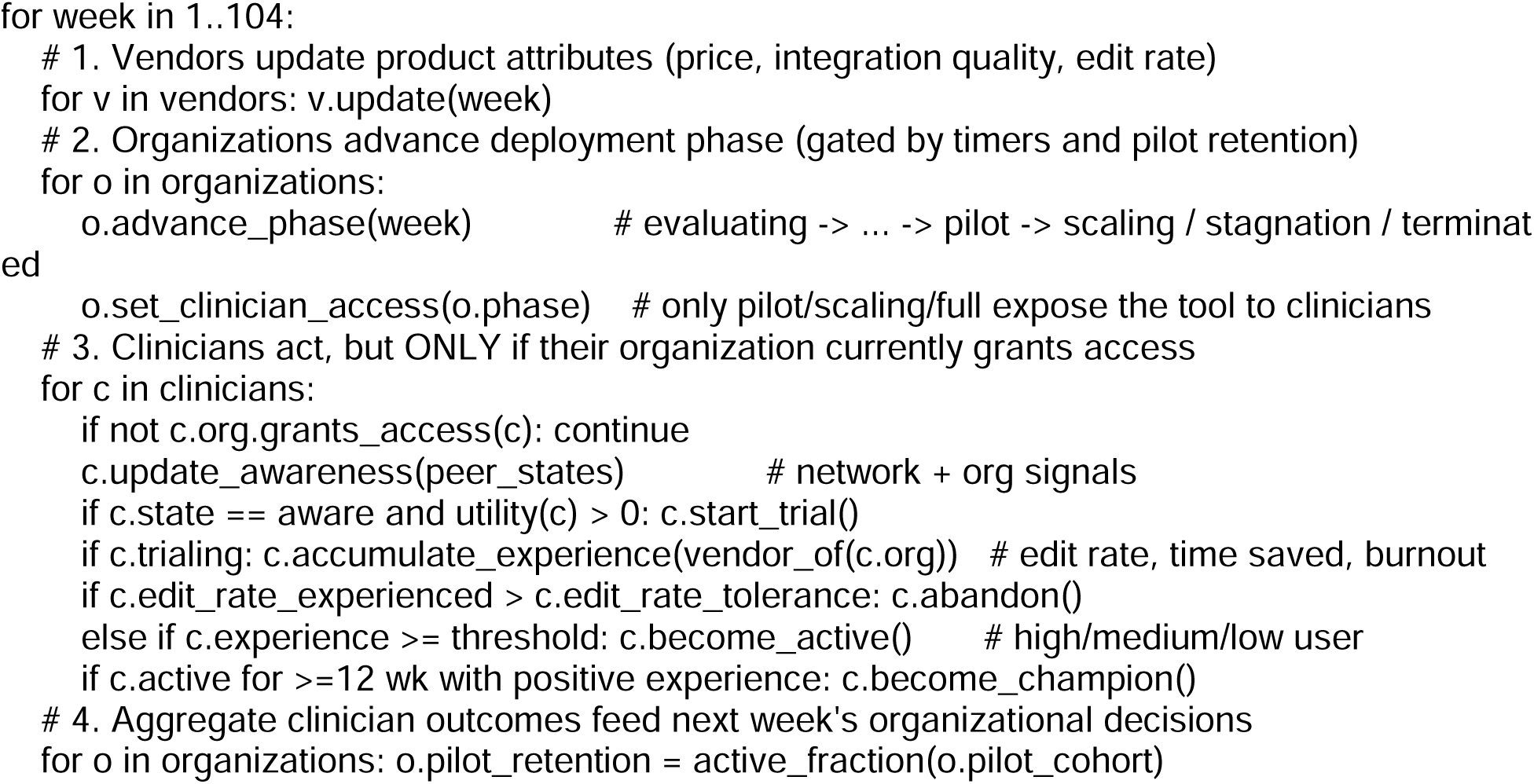

This ordering makes the model’s causal pathways explicit. For example, the **Better EHR Integration** intervention lowers each vendor’s base_edit_rate (step 1); a lower experienced edit rate means fewer trialing clinicians exceed their edit-rate tolerance and abandon, and more accumulate enough positive experience to become active users (step 3); the resulting higher pilot retention makes more organizations clear the scaling threshold rather than entering stagnation or termination (steps 2 and 4), which in turn exposes the tool to more clinicians in subsequent weeks. Adoption and abandonment are thus emergent from the joint clinician-organization-vendor dynamics rather than set directly by any single parameter.

### Calibration and Plausibility Checks

Parameters were anchored to published ranges (**Table S6**): adoption rates of 20-74% [4,5], pilot attrition of 35-45% [5], and usage distributions matching Stanford qualitative findings [13]. We distinguish three uses of empirical data, which should not be conflated: (i) *calibration targets* – published ranges used to set parameter values a priori (**Table S6**); (ii) *pattern plausibility checks* – prespecified qualitative patterns (e.g., psychotype and specialty ordering) evaluated on a separate run set and not used to tune parameters (**Table 2**, **Table S20**, Supplementary Methods S6.2); and (iii) *retrospective comparison* – quantitative goodness-of-fit against published deployment outcomes, computed after the fact (**Table 3**). Importantly, the retrospective comparison is distributional, not site-specific: we did not encode institution-specific characteristics (size, specialty mix, EHR vendor, local governance) of the published sites into matched simulated replicas. Instead, we compare the distribution of simulated outcomes against each published value grouped by reference class, so agreement reflects whether the model’s outcome distribution is consistent with the field’s observed range, not whether it reproduces any individual site. As an additional check, we compared simulated output distributions against published deployment data from seven health systems and three aggregate benchmarks. All cited deployment values were independently re-verified against primary sources before use (Supplementary Methods S8; verification report in the public release). We quantified agreement with formal goodness-of-fit metrics – root-mean-square error (RMSE) and mean absolute error of the model’s predictive median against each observation, 90% predictive-interval coverage, and posterior-predictive tail probabilities – rather than the descriptive range-overlap used in the original submission. Empirical anchors were grouped by reference class (early single-site trajectory; mature single-site final adoption; cross-sectional or randomized ecosystem snapshots), because these reference classes correspond to different model quantities and should not be pooled. Metrics were computed for three model configurations to isolate the effect of successive corrections (cold-start; staggered-only; and the re-calibrated model described below).

### Plateau Re-calibration

The staggered-initialization regime removed the cold-start artifact but raised the steady-state plateau above the cross-sectional and randomized ecosystem benchmarks (final median ∼72%; Results). To bring both the early-time trajectory and the steady-state plateau into agreement with published data simultaneously, we performed a formal calibration over six free parameters: the weekly post-adoption abandonment hazard (base_hazard_rate), the friction threshold triggering abandonment (abandonment_friction_threshold), the pilot-to-scale retention threshold (scale_threshold), the mass placed on seeded deployed phases at initialization (deployed_weight), and the target active-user fractions for organizations seeded into the scaling and full-deployment phases. All other structural parameters – peer influence, champion multiplier, edit-rate tolerance distribution, psychotype proportions, and governance-delay distributions – were held at their prior literature-anchored values. Free-parameter ranges respected the configuration schema (for example, base_hazard_rate is bounded at 5% per week, corresponding to an implausibility ceiling on annual attrition).

The calibration target was the cross-sectional ecosystem plateau (center 0.34, the midpoint of the UCLA randomized 32%, Menlo 35%, and PHTI 20-50% anchors), subject to two constraints: a rising early-time trajectory (not the near-zero cold-start artifact) and an organizational upper tail reaching the motivated single-site range (org-level 90th-percentile final adoption above 50%). We minimized a reference-class-weighted loss combining squared plateau error, an early-time shape penalty, and an upper-tail penalty, searched by Latin-hypercube sampling (100 coarse configurations, four replicates each, then 30 refined configurations at eight replicates), and selected the loss-minimizing configuration. The search established that the achievable plateau floor within schema-valid bounds is approximately 0.40 for the raw ecosystem mean but that the loss-optimal configuration reaches a median final adoption of 0.34 with a valid rising trajectory; the friction threshold and deployed-mass parameters, not the abandonment hazard, are the primary plateau levers. The calibrated configuration (“set G”) was confirmed at full scale (50,000 clinicians, 500 organizations, 10 realizations) and is reported as the primary model for validation (Results; **Table 3**, **Figure 4**). The intervention, factorial, and Sobol analyses below were originally conducted on the original configuration; we re-ran the full six-scenario intervention comparison and the Sobol sensitivity screen natively under set G (the 2^3^ factorial analysis was retained from the original configuration), and report where the calibrated model confirms versus revises the original conclusions (Results, Robustness of Conclusions to Re-calibration).

### Scenario Analysis

We compared baseline dynamics against six intervention scenarios using a paired experimental design (30 realizations per scenario, matched random seeds): (1) Fast-Track Governance (delay reduced to 4 weeks), (2) Champion Program (peer coefficient 0.07 to 0.12), (3) Better EHR Integration (edit rate reduced to 15%), (4) Reduced Resistance proxy (resistant non-adopter fraction set to 0%), (5) High Burnout Crisis, and (6) Combined Intervention (scenarios 1+2+3). Fast-Track Governance is an intentionally aggressive upper-bound scenario (4 weeks vs. mean 18-week governance delay); intervention magnitudes are summarized in **Table S8**. A 2^3^ factorial analysis tested for interactions among the first three factors (Supplementary Methods S9). Effect sizes used Cohen’s d_z_ (paired standardized) with bootstrap confidence intervals (1,000 resamples) [18]; distributional summaries are in **Table S14**.

### Sensitivity Analysis

We performed global sensitivity analysis using Sobol’ indices [19] with 64 base samples (1,152 parameter sets via Saltelli sampling; 2,304 simulations with two replicates each) across eight parameters: abandonment_friction_threshold, governance_delay_mean, edit_rate_tolerance_mean, peer_influence_coefficient, innovation_propensity_alpha, scale_threshold, burnout_reduction_per_use, and champion_influence_multiplier. We report first-order (S_1_) and total-effect (S_T_) indices for final adoption and abandonment rates.

### Emergent Property Analysis

We analyzed 30 independent realizations (50,000 clinicians) to characterize phase transitions, organizational outcome distributions, champion emergence, and specialty-specific adoption heterogeneity.

## Results

### Baseline Adoption Dynamics

Under baseline conditions, the model produced characteristic S-curve adoption trajectories. Mean final active adoption reached 37.6% (SD 2.7%) across 30 paired baseline realizations at full scale (50,000 clinicians). Adoption began with slow uptake as organizations navigated governance approval, followed by acceleration as peer influence compounded, and eventual deceleration as the susceptible population depleted. Inflection timing was multimodal rather than concentrated around a single week: detected inflection points fell into three distinct episodes per run (an early deceleration near week 20, a mid-course acceleration near week 44, and a late deceleration near week 90), so the distribution is better summarized by its median (44 weeks) and interquartile range (20-88 weeks) than by its mean (the mean of 51 weeks falls in a low-density gap between modes and is not representative; **Figure 3A**). Timing also varied between independent run sets (plausibility-check runs centered later, near week 91), reflecting high sensitivity to stochastic initialization and early peer-influence cascades [2]. This path-dependency suggests the model can predict that adoption accelerates but not precisely when, an observation consistent with the unpredictability of real-world tipping points. The ever-tried rate was 55.8% (SD 1.5%), with trial conversion of 49.6% (SD 5.6%) and abandonment of 9.5% (SD 4.5%).

### Intervention Scenarios

Five of six scenarios showed higher adoption relative to baseline (**Table 1**, **Figure 2**). The **Combined Intervention** showed the largest increase (37.6% to 57.5%; +20.0pp; d_z_=5.23). A 2^3^ factorial analysis (**Table S19**, **Figure S4**) revealed that fast governance dominated outcomes (partial η²=0.850), with a significant governance x integration interaction (partial η²=0.046, P=0.001) indicating that streamlined governance amplifies the benefit of lower edit rates; the three-way interaction was not significant (P=0.69). **Fast-Track Governance** produced a large effect (51.4%; +13.9pp; d_z_=2.51), though this reflects an aggressive 78% reduction in governance delay (**Table S8**). **Reduced Resistance** showed a large d_z_ with a modest absolute gain (41.6%; +4.0pp; d_z_=2.08). **Better EHR Integration** (39.4%; +1.8pp; d_z_=0.55) and **Champion Program** (38.6%; +1.0pp; d_z_=0.55) each showed medium effects. **High Burnout Crisis** showed a near-zero effect (38.0%; +0.5pp; d_z_=0.09; CI includes zero), suggesting elevated burnout alone did not reliably accelerate adoption.

**Figure 2.**
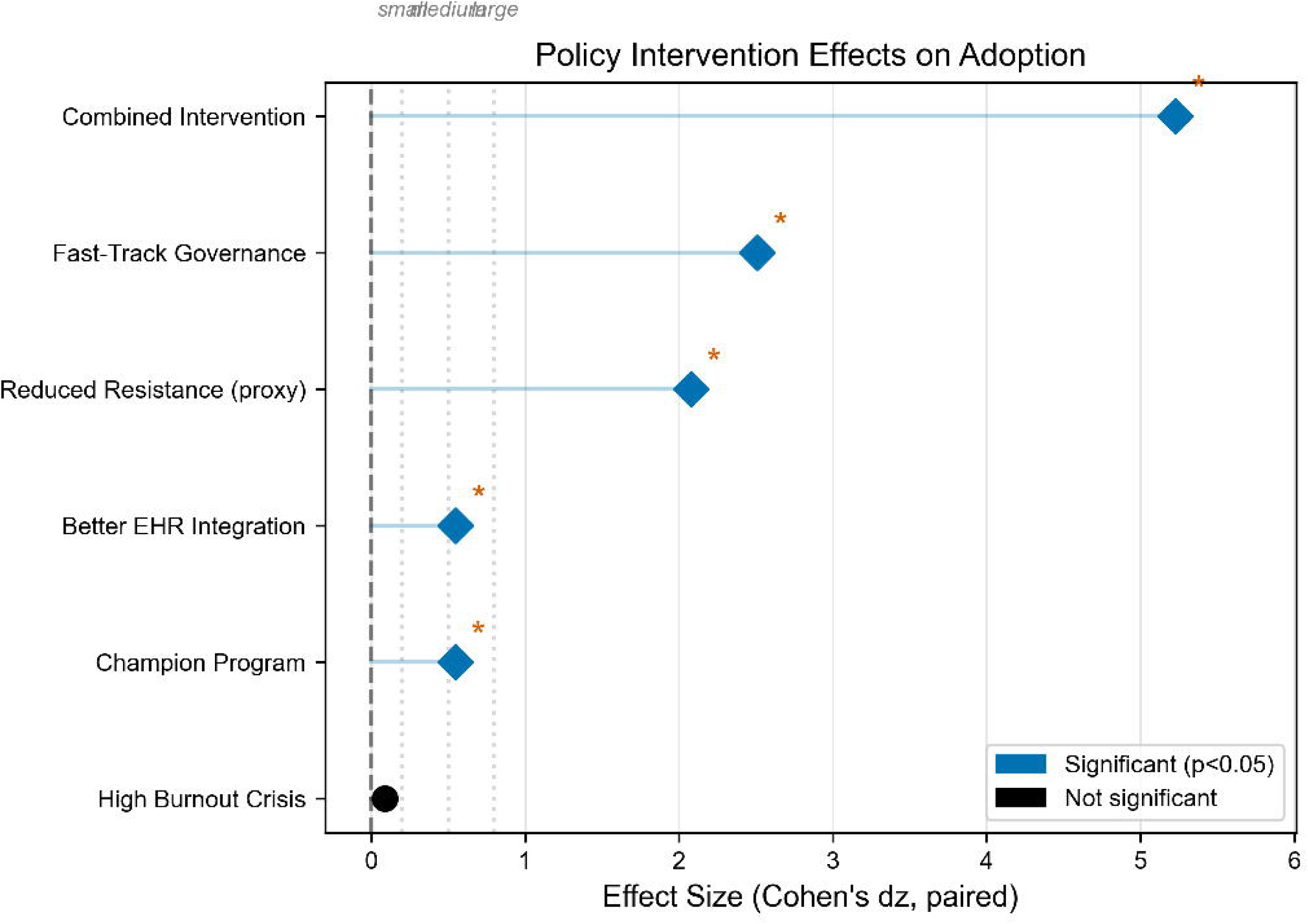
Scenario Comparison Effect Sizes Forest plot showing Cohen’s d_z_ (paired effect size) for six intervention scenarios compared to baseline. Points represent effect estimates (no confidence intervals shown). Vertical dashed lines indicate conventional effect size thresholds: small (0.2), medium (0.5), and large (0.8). Fast-Track Governance shows the largest single-intervention d_z_ (2.51), while the Combined Intervention yields the largest absolute adoption increase (+20.0 pp; d_z_ = 5.23). Reduced Resistance (proxy) shows a large effect (d_z_ = 2.08); Champion Program and Better EHR Integration show medium effects (d_z_ = 0.55). High Burnout Crisis shows a near-zero effect (d_z_ = 0.09).

**Table 1.**
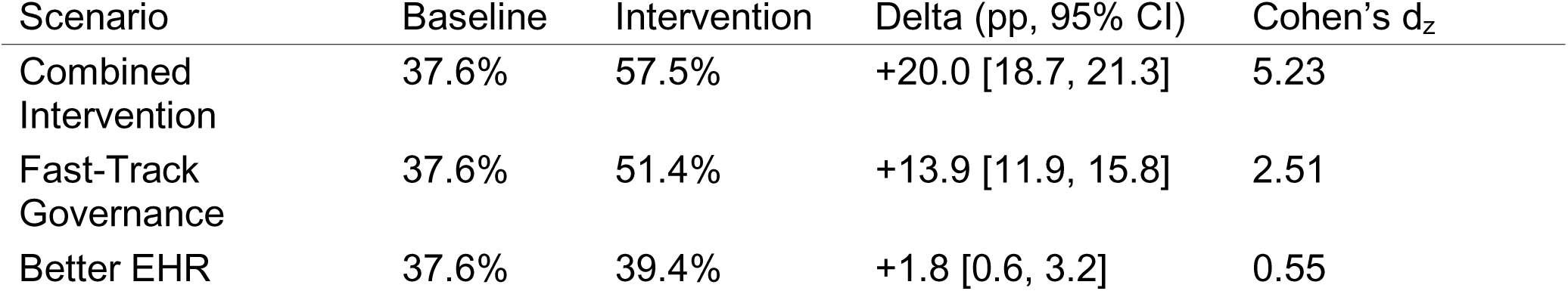
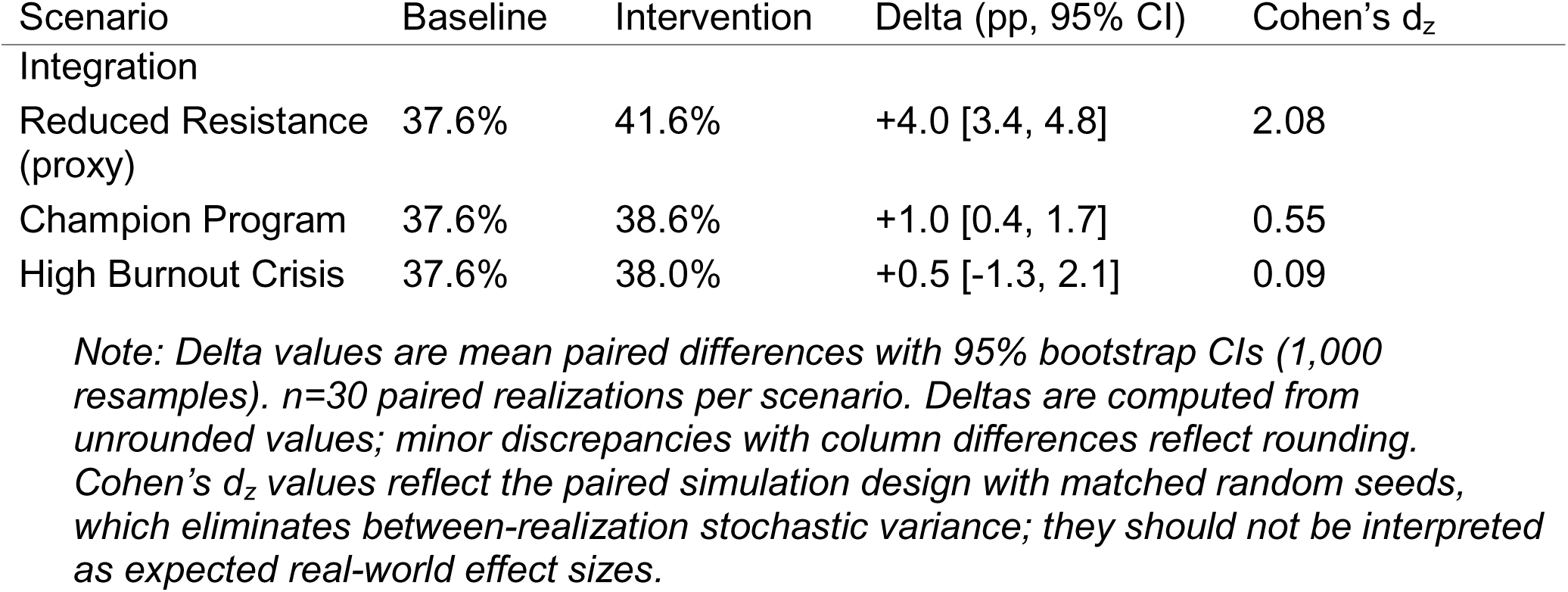
Scenario Comparison Results.

### Sensitivity Analysis

Screening-level Sobol analysis on the original configuration suggested governance delay (S_T_=0.72) and edit-rate tolerance (S_T_=0.34) were the most influential parameters for final adoption (**Figure 5**), with smaller total-effect contributions from burnout reduction, peer influence, and innovation propensity. Confidence intervals were wide and overlapping, so rankings should be interpreted as ordinal rather than precise effect magnitudes. The gap between first-order and total-effect indices indicates non-trivial parameter interactions. This ranking is configuration-dependent: re-running the identical screen under the re-calibrated model changes the leading driver to the abandonment/friction threshold, with edit-rate tolerance again in the top tier and governance delay third (**Robustness of Conclusions to Re-calibration**; **Table S21**).

### Emergent Properties

Adoption trajectories exhibited characteristic inflection-point dynamics (**Figure 3A**), with inflection episodes distributed trimodally (median 44 weeks, IQR 20-88); the previously reported mean of 51 weeks falls between modes and overstates concentration at mid-horizon. Organizations clustered into six outcome categories (**Figure 3B**; original configuration): pilot stagnation (48.9%), success (28.2%), scaling (9.4%), stalled (6.9%), failure (3.4%), and unknown (3.1%). Under re-calibration this distribution re-partitions substantially (pilot stagnation 22.1%, failure 40.5%; **Table 4**), but the aggregate share of organizations in non-success absorbing states is high under both configurations, directionally consistent with reports that many healthcare AI projects remain in pilot or stalled phases [6]; the specific category thresholds are model-defined.

**Figure 3.**
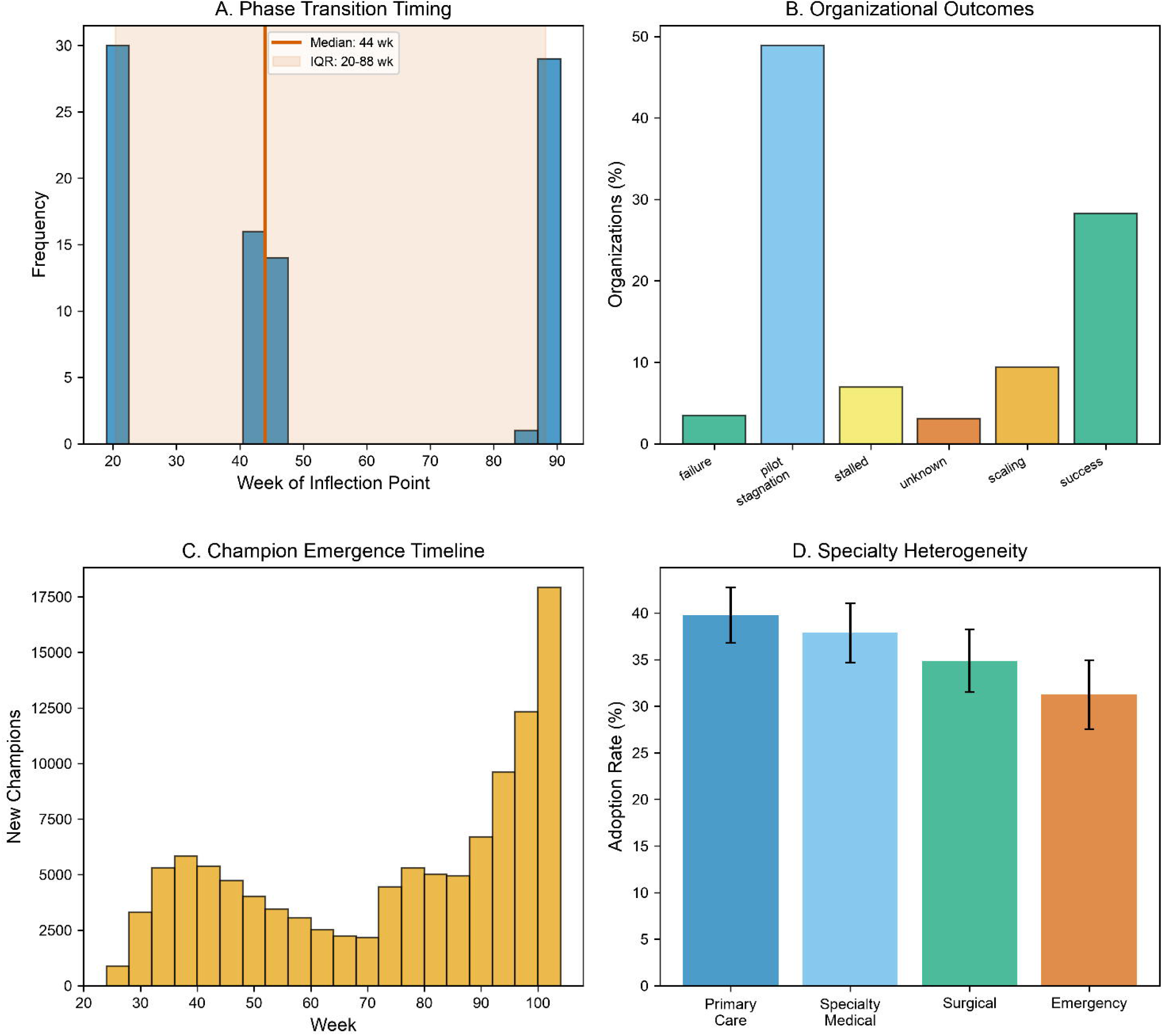
Emergent Properties **(A)** Inflection-point timing. Histogram of detected inflection points across 30 realizations. The distribution is trimodal (an early deceleration near week 20, a mid-course acceleration near week 44, and a late deceleration near week 90) rather than unimodal, so it is summarized by the median (solid line, 44 weeks) and interquartile range (shaded band, 20-88 weeks); the arithmetic mean (∼51 weeks) falls in a low-density gap between modes and is not reported as a central tendency. **(B)** Organizational outcome distribution. Bar chart showing the proportion of organizations in each outcome category at week 104: Pilot stagnation (48.9%, stuck in pilot/early scaling), Failure (3.4%, terminated deployment), Success (28.2%, full deployment achieved), Stalled (6.9%, partial plateau), Scaling (9.4%, active expansion), and Unknown (3.1%, indeterminate). **(C)** Champion emergence timeline. Histogram showing the distribution of weeks when individual champions emerged (defined as sustained users with ≥12 weeks positive experience). Few champions emerged before week 20; emergence rate increased through mid-simulation and declined as the pool of potential champions stabilized. Mean champions at week 104: 3,638 (7.3% of population). **(D)** Specialty-specific adoption rates. Box plots showing final adoption rate by clinical specialty. Primary care showed highest adoption (39.8% ±3.0%), followed by specialty medical (37.9% ±3.2%), surgical (34.9% ±3.4%), and emergency (31.2% ±3.7%). This ordering reflects calibrated documentation burden differentials.

Champion emergence followed a characteristic temporal pattern (**Figure 3C**), with few champions before week 20 (reflecting the 12-week sustained experience requirement) and mean count reaching 3,638 (7.3%) by week 104. No detractors emerged in any realization, likely reflecting model structure rather than real-world absence of vocal critics (see Limitations). Adoption varied systematically by specialty (**Figure 3D**): primary care (39.8%), specialty medical (37.9%), surgical (34.9%), emergency (31.2%), reflecting documentation burden differentials [4].

### Plausibility Checks

Pattern plausibility checks (**Table 2**) showed directional agreement on all six prespecified patterns: correct psychotype and specialty ordering, usage distributions within empirical bands, inflection timing within published ranges, and pilot attrition within the 35-45% acceptance band (35.5%).

**Table 2.** Pattern Plausibility Checks.

| Pattern | Target | Result | Agreement |
| --- | --- | --- | --- |
| Psychotype ordering | EA > Pragmatist > LA > NA | Correct ordering ($\rho=1.0$) | Reproduces |
| Specialty ordering [20] | PC highest; EM lowest | PC > SM > Surg > EM ($\rho=1.0$) | Reproduces |
| Inflection timing | Week 52-104 | Plausibility runs ~91 wk; emergent median 44 wk (IQR 20-88) | Within range |
| Pilot-to-scale attrition | 35-45% | 35.5% | Within range |
| Usage distribution | High 35-50%, Med 20-35%, Low 15-30%, Drop 5-15% | 35.2%, 29.5%, 23.1%, 12.1% | Within range |
| Mature user dropout | 5-15% | 7.7% | Within range |

### Validation Against Published Deployment Data

We compared model output against published deployment data from seven health systems and three aggregate benchmarks (**Table 3**, **Figure 4**) using formal goodness-of-fit metrics, across three successive model configurations: the original cold-start regime, the staggered-initialization regime, and the re-calibrated model (set G; Methods). All cited deployment values were independently re-verified against primary sources before use, and anchors were grouped by reference class (early single-site trajectory; mature single-site final adoption; cross-sectional or randomized ecosystem snapshots) because they correspond to different model quantities and must not be pooled. Metrics were computed over 50 realizations per configuration with 1,000-resample bootstrap confidence intervals on each RMSE.

**Figure 4.**
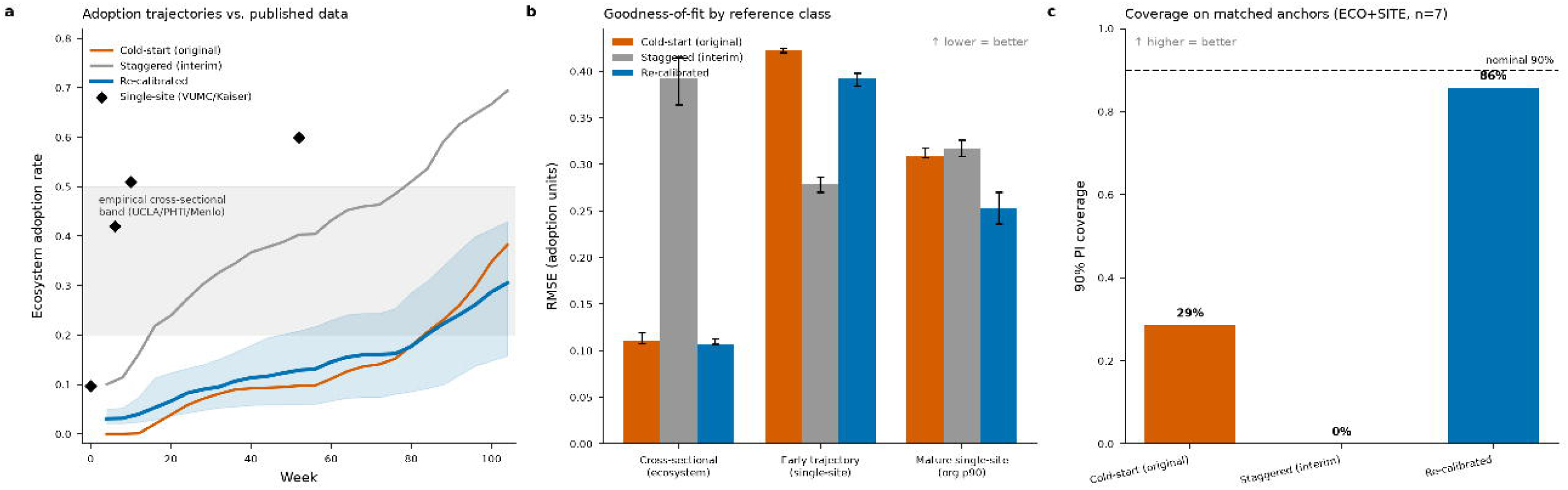
Validation Against Published Deployment Data Across Model Configurations Three-panel comparison of the model against published deployment data under three successive configurations: the original cold-start regime, the staggered-initialization regime, and the re-calibrated model (set G). **(A)** Median ecosystem adoption trajectories for each configuration, with the empirical cross-sectional band (20-50%; UCLA/PHTI/Menlo) shaded and single-site trajectory points (VUMC, Kaiser; diamonds) overlaid; the re-calibrated trajectory (shaded 5th-95th percentile envelope) rises immediately and plateaus within the empirical band, whereas cold-start stays near zero for ∼40 weeks and staggered-only overshoots. **(B)** Root-mean-square error by empirical reference class (cross-sectional ecosystem; early single-site trajectory; mature single-site final adoption), with 95% bootstrap confidence intervals; the re-calibrated model achieves the best fit on the cross-sectional and mature single-site classes. **(C)** 90% predictive-interval coverage on the two reference classes whose model analogs properly match the empirical quantities (ecosystem + mature single-site; seven anchors): 29% cold-start, 0% staggered-only, 86% re-calibrated, against the nominal 90% (dashed line). Fifty realizations per configuration. See Supplementary Methods S8 and main-text Table 3.

**Figure 5.**
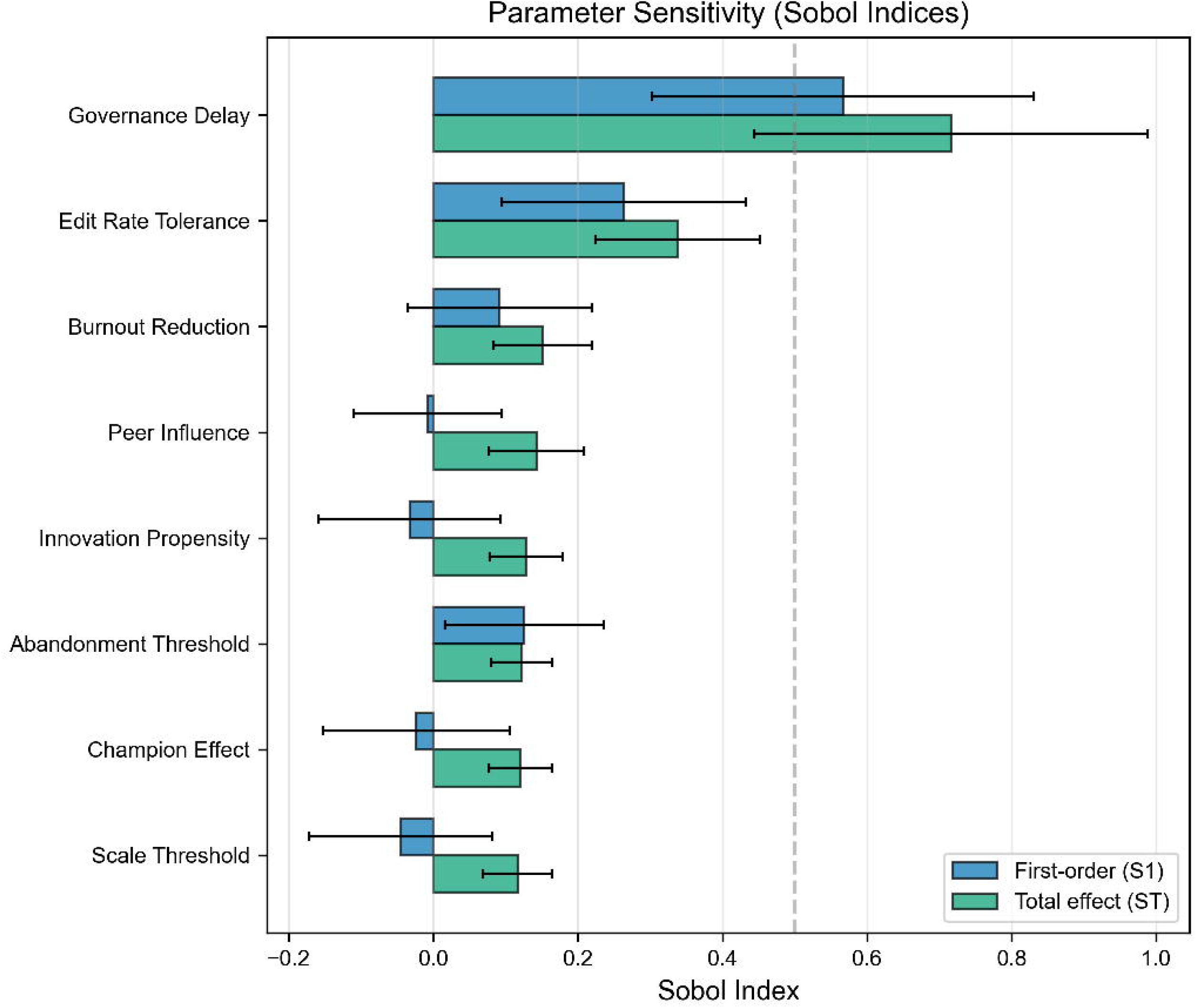
Global Sensitivity Analysis Tornado plot showing Sobol sensitivity indices for final adoption rate. Blue bars indicate first-order indices (S_1_, variance from parameter alone); green bars indicate total-effect indices (S_T_, variance including all interactions). Error bars represent 95% bootstrap confidence intervals. Parameters are sorted by S_T_ in descending order; rankings are screening-level with overlapping uncertainty. Governance delay and edit-rate tolerance show the largest total effects in this analysis (original configuration). The gap between S_1_ and S_T_ indicates parameter interactions.

Under cold-start initialization, all organizations begin in a not-started state, so the modeled ecosystem produced near-zero adoption for the first ∼10 weeks: every early single-site trajectory observation fell outside the model’s 90% predictive interval (trajectory RMSE 0.42, coverage 0/4), and no mature single-site final-adoption value was covered (RMSE 0.31, coverage 0/3). The cold-start regime did fit the lower cross-sectional and randomized ecosystem anchors (RMSE 0.11, coverage 2/4), but for the wrong reason – the artifactual suppression of early adoption depressed the ecosystem mean rather than reproducing the underlying dynamics. Staggered initialization removed the cold-start artifact and improved trajectory fit (RMSE 0.28) but overshot the steady-state plateau (final median 72.1%), worsening the ecosystem-snapshot fit (RMSE 0.39, coverage 0/4) and failing to cover any mature single-site anchor.

The re-calibrated model resolved this tradeoff. Jointly calibrating the plateau while preserving a rising early-time trajectory brought the ecosystem snapshots back into agreement legitimately (RMSE 0.11, coverage 3/4) while simultaneously achieving the best fit to the mature single-site anchors (RMSE 0.25, coverage 3/3) – the only configuration to cover all three single-site final-adoption values. Pooling across the two reference classes that the ecosystem mean and organization-level distribution properly map to (cross-sectional plus mature single-site; seven anchors), the re-calibrated model’s 90% predictive intervals covered 6/7 (86%) of observations, near the nominal 90%, versus 2/7 (29%) for cold-start and 0/7 for staggered-only (**Figure 4C**). At full scale (50,000 clinicians, 500 organizations, 10 realizations) the re-calibrated model reproduced this plateau (final median 36.4%, mean 33.6%, SD 11.4%), with 8 of 10 realizations inside the empirical cross-sectional band (20-50%). The early single-site trajectory anchors (VUMC 42-51%, Kaiser 60% within the first year) remain outside every configuration’s ecosystem-level interval, because these are single-site quantities measured within motivated early-adopter organizations and their model analog is organization-level, not ecosystem-mean, adoption; the mature single-site reference class captures that comparison (org-level 90th-percentile final adoption), and only the re-calibrated model covers it. This comparison is post-hoc and subject to selection bias; detailed interpretation is in Supplementary S8 and S10.

**Table 3.**
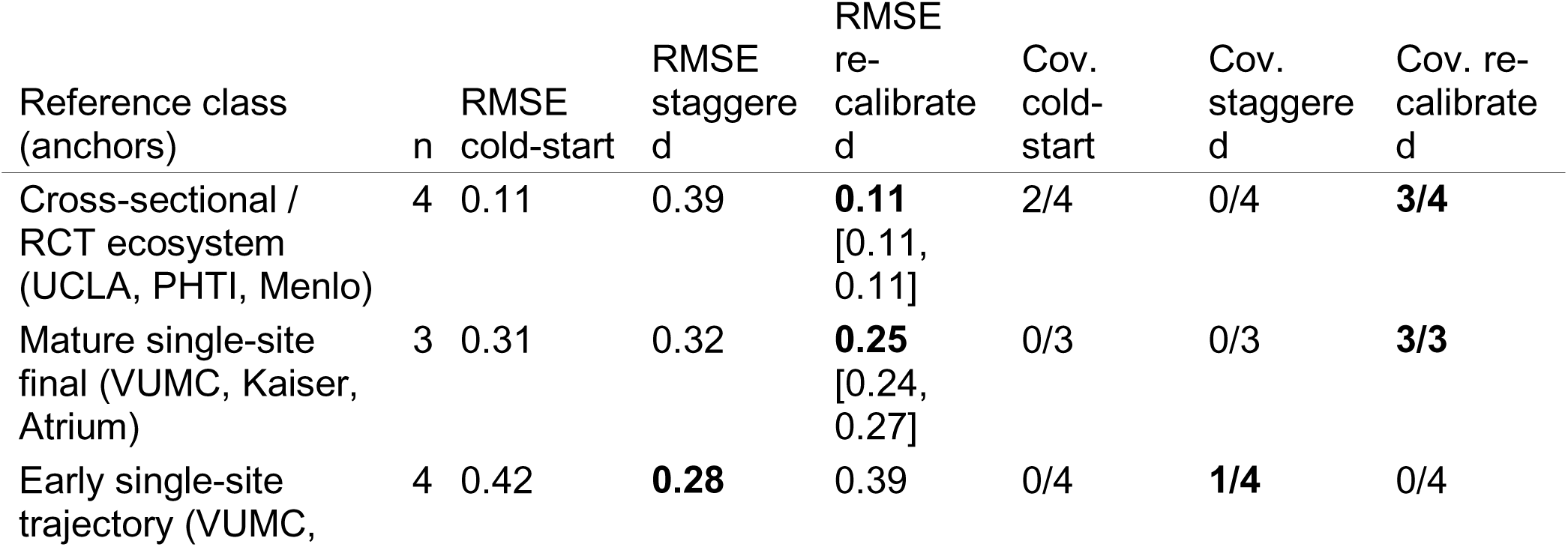
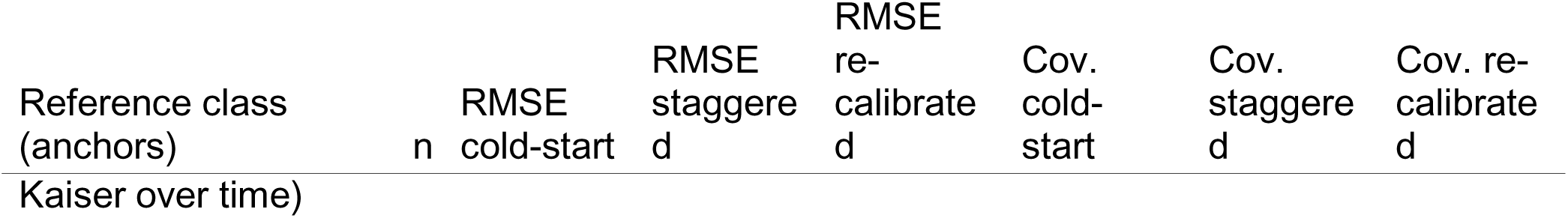
Quantitative Validation Against Published Deployment Data. Goodness-of-fit metrics by empirical reference class, across three model configurations (50 realizations each). RMSE is of the model’s predictive median against each observation (adoption-rate units; 95% bootstrap CI in brackets); coverage is the fraction of observations inside the model’s 90% predictive interval (higher is better; an ideal nominal-90% interval covers ∼0.9). Bold marks the best-fitting configuration per row.

Pooled 90% predictive-interval coverage on the two matched reference classes (cross-sectional ecosystem + mature single-site; seven anchors), whose model analogs – the ecosystem mean and the organization-level distribution – properly match the empirical quantities: **cold-start 2/7 (29%), staggered 0/7 (0%), re-calibrated 6/7 (86%)** (**Figure 4C**).

*The re-calibration was targeted at the adoption level and trajectory shape; the six free parameters were tuned to these deployment reference classes (Methods, Plateau Re-calibration). The unverified Ambience retention figure used in the original submission was removed; the retention dimension is carried by the PHTI and Stanford/Atrium usage anchors. Specialty ordering (rho = 1.0 vs. consensus PC>SM>Surg>EM) and the Stanford usage distribution were matched under all configurations*.

### Robustness of Conclusions to Re-calibration

The intervention, factorial, and Sobol analyses above were conducted on the original configuration. To test whether their qualitative conclusions survive, we re-ran the full six-scenario comparison and the Sobol screen natively under the re-calibrated model (set G). All six intervention scenarios were re-run as paired contrasts (30 realizations each) at the representative validation scale (8,000 clinicians, 160 organizations), whose baseline adoption (34.0%, SD 8.8%) matches the confirmed full-scale baseline (33.6%; §Full-Scale Confirmation) more closely than the smaller pilot scale used during calibration development. The re-calibrated intervention effects are summarized in **Table 5**.

Three points emerged, two of which revise conclusions from the original configuration. First, under re-calibration the intervention effects are considerably smaller and the standardized effect sizes are consistent with genuine, larger between-realization variability (baseline SD 8.8% vs. 2.7% originally); most single-lever effects have paired-d_z_ confidence intervals that include zero. Second, and most consequentially, **the combined intervention no longer exceeds every single lever**: at the matched scale, fast-track governance alone produced the largest gain (+4.6pp) while the combined intervention produced +3.2pp. We verified that this is not a sampling artifact — two independent runs (20 and 30 realizations) both showed the combined effect at or below fast-track governance, and the ordering is a consequence of scale: at the smaller pilot scale (4,000/80, baseline 31.8%) the combined effect was +13.5pp and clearly largest, but that scale’s baseline is an outlier and effects roughly halve as the system approaches its full-scale operating point (**Figure 6A**). Because the manuscript scale is 500 organizations, the smaller-scale figures overstate intervention leverage. Third, one lever is robust: reduced resistance (redistributing the resistor psychotype to pragmatists) produced a small but highly consistent effect (+1.7pp; d_z_=0.90, 95% CI [0.57, 1.45]) because it acts uniformly on every clinician rather than through high-variance organizational gates. Fast-track governance retained the largest mean effect but with a confidence interval spanning zero (d_z_=0.32 [-0.04, 0.80]), so it is best described as the largest *structural* lever rather than a statistically reliable one at this scale. The champion program, a modest positive in the original, remained null under re-calibration (+0.4pp; d_z_=0.19 [-0.17, 0.64]).

**Figure 6.**
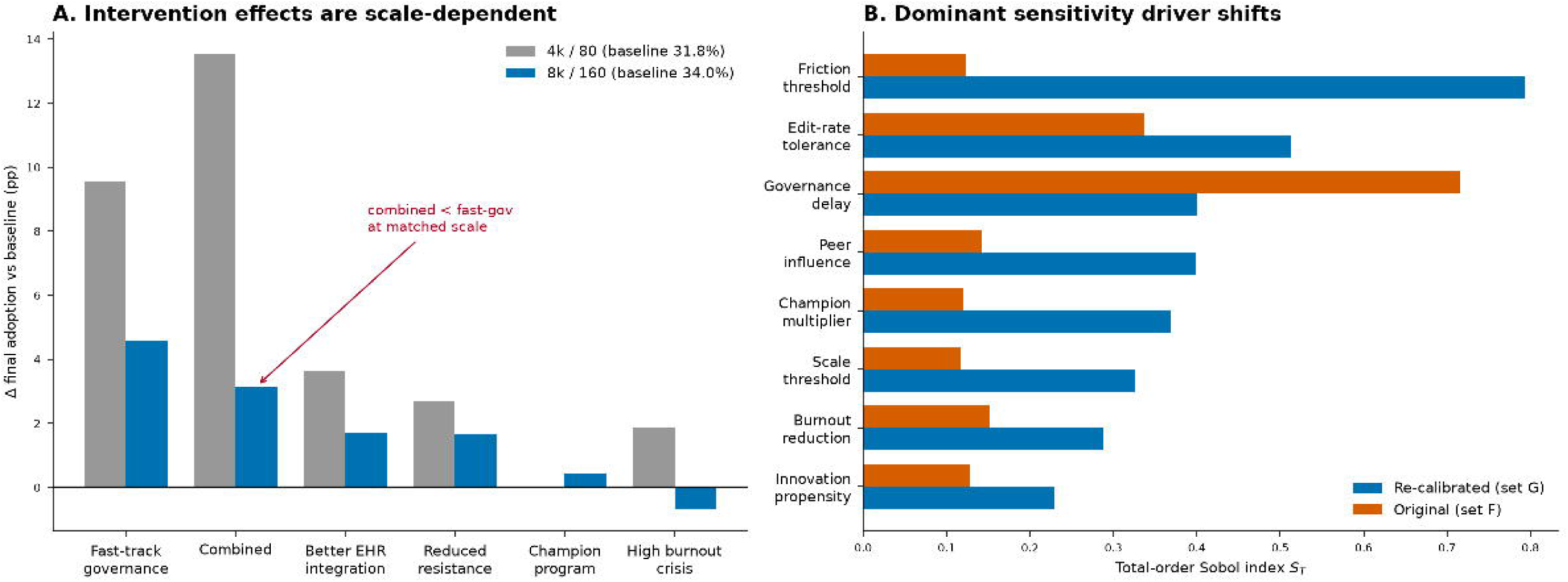
Native Re-run of Interventions and Sensitivity Under the Re-calibrated Model Results of re-running the full scenario and sensitivity analyses natively under the re-calibrated model (set G). **(A)** Intervention effects (change in final adoption relative to baseline, in percentage points) for all six scenarios at two population scales: the small pilot scale (4,000 clinicians / 80 organizations; baseline 31.8%) and the representative validation scale (8,000 / 160; baseline 34.0%, matching the confirmed full-scale baseline of 33.6%). Effects roughly halve at the larger scale, and the combined intervention falls below fast-track governance alone at the matched scale (annotated), so the “combined exceeds every single lever” ordering is a small-scale artifact. **(B)** Total-order Sobol indices (S_T_) for final adoption under the re-calibrated model (set G) versus the original configuration; the dominant driver shifts from governance delay (original) to the abandonment/friction threshold (re-calibrated), with edit-rate tolerance remaining a top-tier driver across both. Sobol screen: 64 base samples, 1,152 parameter sets; intervention scenarios: 30 paired realizations at 8,000/160. See Results (Robustness of Conclusions to Re-calibration), main-text Table 5, and Supplementary Table S21.

A Sobol screen re-run natively under set G (identical eight-parameter design; 64 base samples, 1,152 parameter sets; **Table S21**) likewise revised the sensitivity ranking. In the original configuration governance delay dominated (S_T_=0.72); under re-calibration the abandonment/friction threshold became the single most influential parameter (S_T_=0.79), with edit-rate tolerance second (S_T_=0.51) and governance delay third (S_T_=0.40) (**Figure 6B**). This is mechanistically consistent with the calibration itself, which made clinician friction and churn the primary determinants of the steady-state plateau. Edit-rate tolerance remained a top-tier driver across both configurations; governance delay remained influential but is no longer the leading parameter once the model is calibrated to real adoption levels.

Organizational outcome clustering also shifted under re-calibration (**Table 4**). Raising the abandonment hazard and pilot-to-scale threshold to lower the plateau necessarily reclassifies organizations that previously stagnated as terminating: the re-calibrated model shows organizational failure at 40.5% (originally 3.4%) and pilot stagnation at 22.1% (originally 48.9%), with success at 23.4% (originally 28.2%). The original conclusion that pilot stagnation is the single most prevalent outcome therefore does not survive re-calibration; the defensible restatement is that pilot stagnation and outright failure are both prominent absorbing states (together ∼63%), consistent with reports of stalled healthcare AI projects but with the specific partition between stagnation and failure being model-defined and threshold-sensitive.

**Table 4.** Organizational Outcome Distribution: Original vs. Re-calibrated.

| Outcome | Original (%) | Re-calibrated (%) |
| --- | --- | --- |
| Success (full deployment) | 28.2 | 23.4 |
| Scaling | 9.4 | 9.1 |
| Pilot stagnation | 48.9 | 22.1 |
| Stalled (pre-pilot) | 6.9 | 2.9 |
| Failure (terminated) | 3.4 | 40.5 |
| Unknown | 3.1 | 2.0 |

**Table 5.** Intervention Effects Under Re-calibration (Set G; 8,000 clinicians, 160 organizations, 30 paired realizations)

| Scenario | Mean adoption (%) | $\Delta$ vs baseline (pp) | Cohen's $d_z$ | $d_z$ 95% CI |
| --- | --- | --- | --- | --- |
| Re-calibrated baseline | 34.0 | — | — | SD 8.8% |
| Fast-Track Governance | 38.5 | +4.6 | 0.32 | [-0.04, 0.80] |
| Combined Intervention | 37.1 | +3.2 | 0.20 | [-0.16, 0.58] |
| Better EHR Integration | 35.7 | +1.7 | 0.24 | [-0.11, 0.62] |
| Reduced Resistance | 35.6 | +1.7 | 0.90 | [0.57, 1.45] |
| Champion Program | 34.4 | +0.4 | 0.19 | [-0.17, 0.64] |
| High Burnout Crisis | 33.3 | -0.7 | -0.05 | [-0.41, 0.35] |

In summary, re-running the full scenario and sensitivity analyses natively under the calibrated model both confirmed and revised the original conclusions. Confirmed: governance remains an influential upstream lever, the largest single structural intervention; factorial analysis (original configuration) identified a significant governance x integration interaction; and a large share of organizations settle into non-success absorbing states. Revised: the combined intervention does not reliably exceed the best single lever once the model is calibrated and evaluated at scale; the leading sensitivity driver shifts from governance delay to clinician friction/edit-rate tolerance; and the organizational-outcome partition moves from pilot-stagnation-dominant to a mix of stagnation and failure. These revisions strengthen rather than weaken the paper’s central methodological claim — that calibrating the model to real adoption levels materially changes which levers appear to matter — and reinforce that the framework is best used for comparative hypothesis generation rather than point ranking of interventions.

## Discussion

This study presents an agent-based modeling framework for healthcare AI adoption applied to ambient clinical documentation. Three principal findings emerge. First, governance delay is an influential upstream lever and the largest single *structural* intervention: in the original configuration it produced the largest modeled effect (+13.9pp, d_z_=2.51) and ranked first in the Sobol screen (S_T_=0.72). Because governance is an explicit upstream gate, this leverage is structural by design and should be interpreted as influence within model architecture rather than evidence that governance is the most important real-world lever; the fast-track result is an upper-bound estimate (**Table S8**). This leadership is also configuration-dependent: when the model is re-calibrated to real adoption levels and the analyses are re-run natively, governance delay drops to third in the sensitivity ranking behind clinician friction/edit-rate tolerance, and its intervention effect, while still the largest single lever, is no longer statistically distinguishable from zero at full operating scale (Results, Robustness of Conclusions to Re-calibration). Second, a 2^3^ factorial analysis revealed that fast governance dominated outcomes (partial η²=0.850), with a significant governance x integration interaction (P=0.001) indicating that streamlined governance amplifies the benefit of lower edit rates. Third, a large share of simulated organizations settled into non-success absorbing states rather than reaching full deployment; the specific partition of that share between pilot stagnation and outright failure is model-defined and shifts with calibration (pilot stagnation 48.9% in the original configuration versus 22.1% under re-calibration, with organizational failure rising correspondingly), but the aggregate finding that most organizations do not reach sustained full deployment is directionally consistent with reports of stalled healthcare AI projects [6] and robust across configurations.

The model indicates several dynamics not well-characterized in existing literature. Prior studies have documented individual clinician adoption variability (20-74% [2,4,5]), but the organizational-level distribution – with success, pilot stagnation, and failure as distinct attractor states – represents a model prediction. The prominence of non-success absorbing states (pilot stagnation plus failure together the majority of organizations under both configurations) suggests organization-level factors may explain as much variance as individual characteristics. Edit-rate tolerance dominated abandonment sensitivity (S_T_=0.88), consistent with evidence that editing burden drives discontinuation [13]. The Champion Program yielded only a modest gain in the original configuration (d_z_=0.55) and a null effect under re-calibration (d_z_=-0.01); the 2× champion multiplier is already an optimistic assumption relative to empirical estimates (∼1.2-1.5x), and the absence of modeled detractors likely inflates peer-support estimates – so the weakened champion effect under the higher-churn re-calibrated regime should be read as a caution against over-crediting peer-amplification levers in isolation. Despite calibration to empirical burnout prevalence [16] and reduction estimates [17], the High Burnout Crisis showed near-zero adoption effect, suggesting elevated burnout alone does not reliably accelerate adoption.

Agent-based modeling offers advantages over alternative approaches for this domain. Traditional diffusion models (Bass, logistic growth) assume population homogeneity and fit a single aggregate curve; they can describe that adoption follows an S-shape but cannot represent psychotype-specific decision rules, the organizational gating that precedes clinician access, or the two-sided coupling between them. System dynamics models aggregate individual decisions into stock-flow structures, obscuring network effects and emergent organizational outcomes [22]. The added value of the present framework is precisely in what those approaches cannot produce: it makes the *mechanism* connecting an intervention to an outcome explicit and testable, and it generates emergent, distributional predictions (the spread of organizational trajectories, the multimodal timing of inflection points, the partition of organizations into absorbing states) rather than a single fitted curve. The re-calibration exercise illustrates why this matters: because the levers act through different mechanisms (governance through an upstream timing gate, edit-rate tolerance and reduced resistance through per-clinician friction), calibrating the model to realistic adoption levels re-ordered which levers appear influential – a dependency that an aggregate curve-fit cannot surface. Our approach aligns with calls for complexity-aware methods in health technology assessment, including the NASSS framework’s emphasis on multi-level adoption dynamics [23]. Extended methodological comparison is in Supplementary S10.

The framework is not specific to ambient documentation. Its transferable elements – heterogeneous clinician decision agents, an organizational deployment-phase gate, vendor attributes, a peer-influence network, and the weekly cross-level update loop – are common to the adoption of many clinical technologies (for example, clinical decision support, diagnostic AI, or patient-facing tools), and re-targeting the model requires substituting the technology-specific inputs (the relevant friction and benefit measures, the governance timeline, and vendor characteristics) rather than restructuring the model. Ambient documentation was chosen as the calibration case because it is one of the first AI technologies with enough published multi-site deployment data to anchor and test such a model; the same architecture could be re-calibrated as comparable data accrue for other technologies. What would not transfer without re-estimation are the numerical calibration targets and the intervention magnitudes, which are documentation-specific.

These findings are hypothesis-generating rather than prescriptive. They suggest empirical studies could compare governance process changes of varying magnitude, measure edit-burden dynamics longitudinally, and test whether peer programs amplify adoption when paired with integration improvements. The model does not distinguish friction from careful safety review versus poor integration; implementation studies should pair integration metrics with safety monitoring [21]. Organizational heterogeneity suggests uniform policies may have variable effects across health systems.

Several limitations warrant consideration. Although the re-calibrated model reproduces published adoption levels and trajectories across reference classes, it should be interpreted as a comparative-scenario framework rather than a point-forecasting tool, given its wide between-realization variability and reliance on a small, post-hoc anchor set. Plausibility checks show directional agreement but are not fully independent from calibration priors. Sobol indices have wide confidence intervals at N=64 base samples, and rankings are not statistically distinguishable. No detractors emerged, reflecting a modeling choice that routes dissatisfied users to abandonment rather than persistent vocal criticism; this makes peer influence systematically one-directional, inflating net social influence effects and baseline adoption trajectories. The peer influence parameter itself derives from prescribing network studies [7], a different decision domain than technology adoption, adding further uncertainty to network-mediated results. The model’s original cold-start regime initialized all organizations in a not-started state, which suppressed ecosystem adoption for the first ∼10 weeks and prevented early-time comparison with published sites observed mid-rollout. We addressed this in two steps: a staggered-initialization regime calibrated to verified governance-to-deployment timelines, which removed the artifact but overshot the steady-state plateau; and a formal six-parameter re-calibration that brings both the early-time trajectory and the steady-state plateau into agreement with published data, covering 86% of the properly reference-class-matched anchors at the nominal 90% level (Results; **Table 3**, **Figure 4**). We regard the re-calibrated model as a better-supported configuration but not a validated point-forecasting tool: the calibration targets a small number of published reference classes, the anchor set is post-hoc and subject to selection bias, and the re-calibrated model exhibits wide between-realization variability (final-adoption SD 11.4% at full scale) that bounds any point prediction. We report the re-calibrated model as the primary configuration for validation and re-ran the intervention comparison and the Sobol sensitivity screen natively under it (Results, Robustness of Conclusions to Re-calibration); the 2^3^ factorial analysis is retained from the original configuration and its central interaction was not re-estimated under re-calibration, which is a residual limitation. The small-world network topology was not varied in sensitivity analysis; adoption dynamics may differ under alternative structures. The model does not represent patient outcomes, quality metrics, or cost-effectiveness. Psychotype proportions (15/40/30/15%) are substantially adapted from Rogers’ original categories (see Supplementary S1.2), and several calibration targets draw on proxy measures that may not transfer directly to ambient documentation adoption. Additional limitations regarding parameter uncertainty, evidence heterogeneity, network structure, and generalizability are discussed in Supplementary S10.

Longitudinal multi-site studies tracking individual and organizational adoption trajectories would enable validation of predicted outcome distributions. Multi-site comparative studies could estimate parameters that our sensitivity analysis identified as influential but poorly characterized. Extending the model to capture detractor dynamics, vendor co-evolution, and learning-curve effects [14] could improve realism. The framework could support prospective exploration of implementation strategies before deployment, allowing health systems to prioritize empirical evaluations of high-impact candidates.

## Conclusions

Agent-based modeling offers a structured framework for understanding healthcare AI adoption dynamics. Applied to ambient clinical documentation, one of the first widely deployed AI technologies in clinical practice, the model highlights governance processes as a key bottleneck within modeled conditions, reveals significant intervention interactions through factorial analysis, and predicts that most organizations settle into non-success absorbing states (pilot stagnation and failure) as an emergent system property. After correcting an initialization artifact and re-calibrating, the model reproduces both the early-time adoption trajectory and the steady-state plateau of published multi-site deployment data. Re-running the full intervention and sensitivity analyses natively under the calibrated model confirmed some conclusions and revised others: governance remains influential and non-success absorbing states remain prominent, but the combined intervention no longer reliably exceeds the best single lever at operating scale and the leading sensitivity driver shifts from governance to clinician friction. That the calibration changes which levers appear to matter is itself the central methodological lesson. These findings are hypothesis-generating rather than prescriptive. The model does not assess impacts on safety oversight or safety outcomes. As healthcare AI expands into diagnostic, therapeutic, and administrative domains, quantitative frameworks for anticipating adoption trajectories and guiding empirical study design may become increasingly valuable.

## Supporting information

Supplementary Results

## Ethics Approval

This study is a computational simulation and did not involve human participants, human tissue, or identifiable patient data; no institutional review board approval or informed consent was required. All empirical values used to parameterize and validate the model were obtained from previously published, publicly available aggregate sources, which are cited in the manuscript and Supplementary Methods. All relevant ethical guidelines for research not involving human or animal subjects have been followed.

## Data Availability

The model produces synthetic (simulated) data only; no patient data or other human-subjects data were used. Simulation code, analysis and calibration scripts, configuration files, and the empirical-source verification report will be deposited in a public repository upon posting. The published aggregate deployment values used for validation are available in the cited primary sources.

## CRediT authorship contribution statement

Matthew G. Crowson: Conceptualization, Methodology, Software, Validation, Formal Analysis, Investigation, Resources, Writing -Original Draft, Writing -Review & Editing, Visualization, Project Administration.

## Acknowledgments

None.

## Supplementary Figures

**Figure S1.**
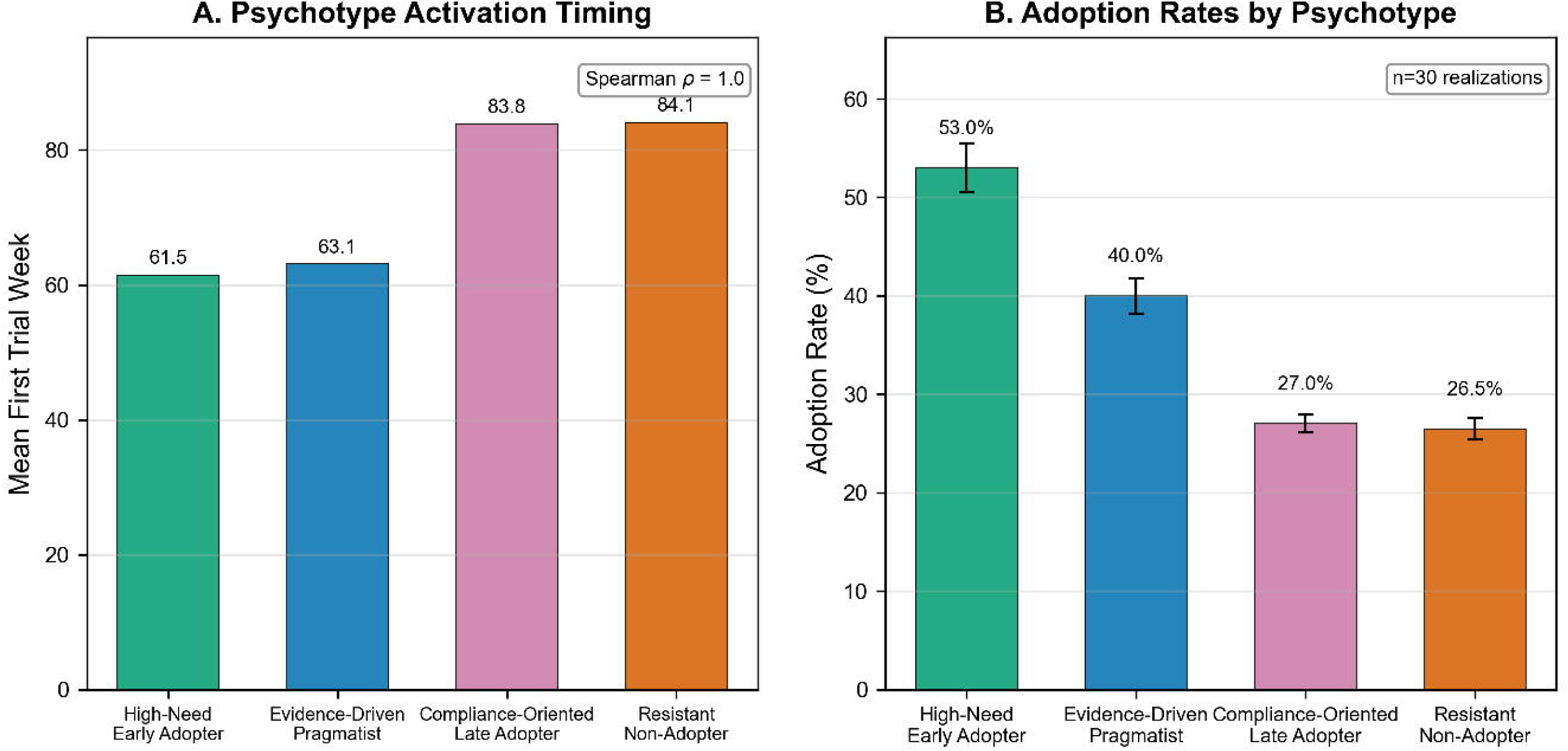
Psychotype validation analysis. **(A)** Psychotype activation timing showing first trial week by adopter type: High-Need Early Adopters activate earliest, followed by Evidence-Driven Pragmatists, Compliance-Oriented Late Adopters, and Resistant Non-Adopters (correct rank ordering across 4 categories), consistent with Rogers diffusion theory. **(B)** Adoption rates by psychotype showing the expected ordering from highest (High-Need Early Adopter) to lowest (Resistant Non-Adopter) adoption.

**Figure S2.**
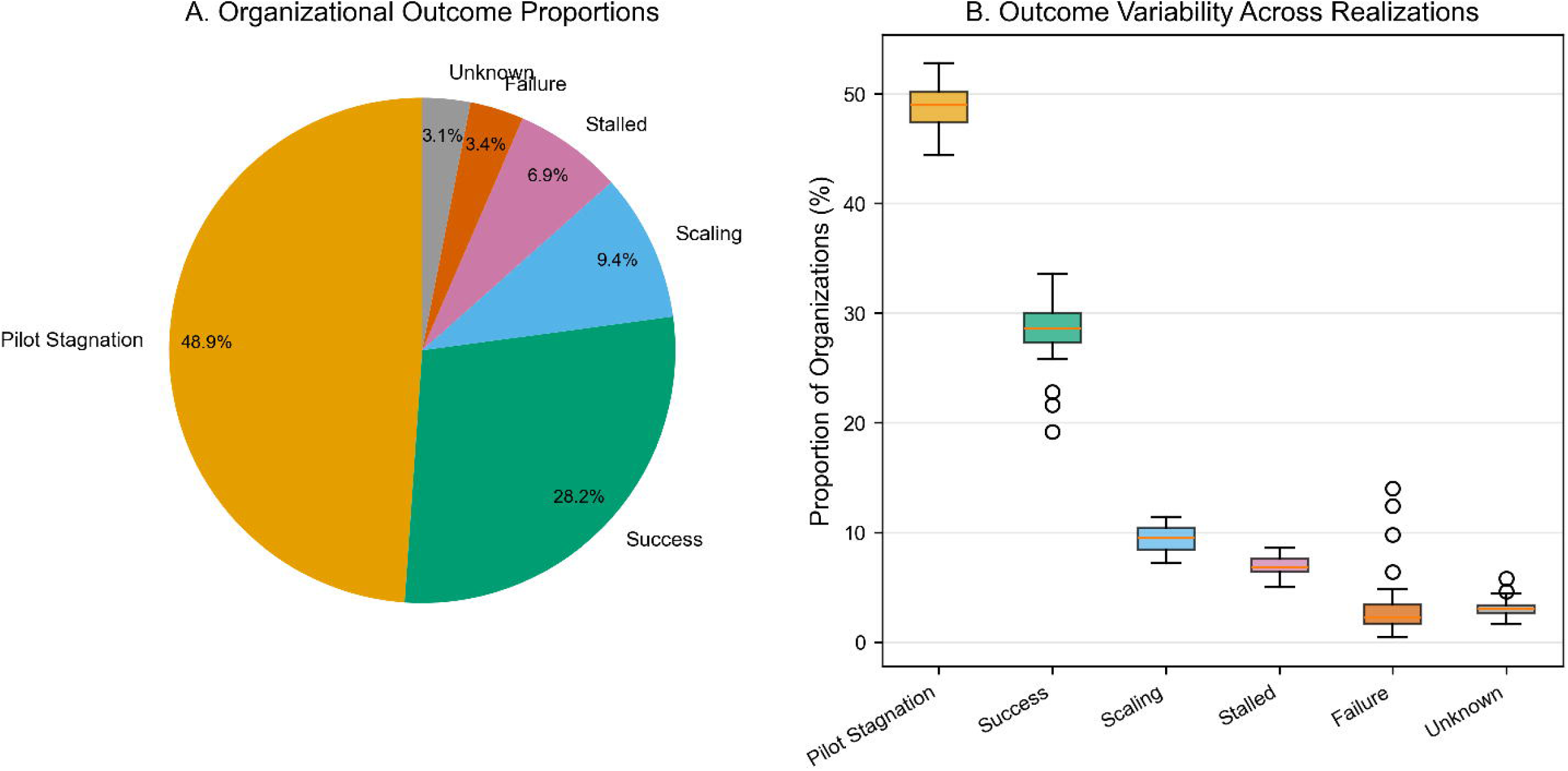
Organizational outcome distribution across deployment phases at week 104. **(A)** Pie chart showing proportions: pilot stagnation (48.9%), success (28.2%), scaling (9.4%), stalled (6.9%), failure (3.4%), and unknown (3.1%). **(B)** Box plots showing outcome rate variability across 30 realizations.

**Figure S3.**
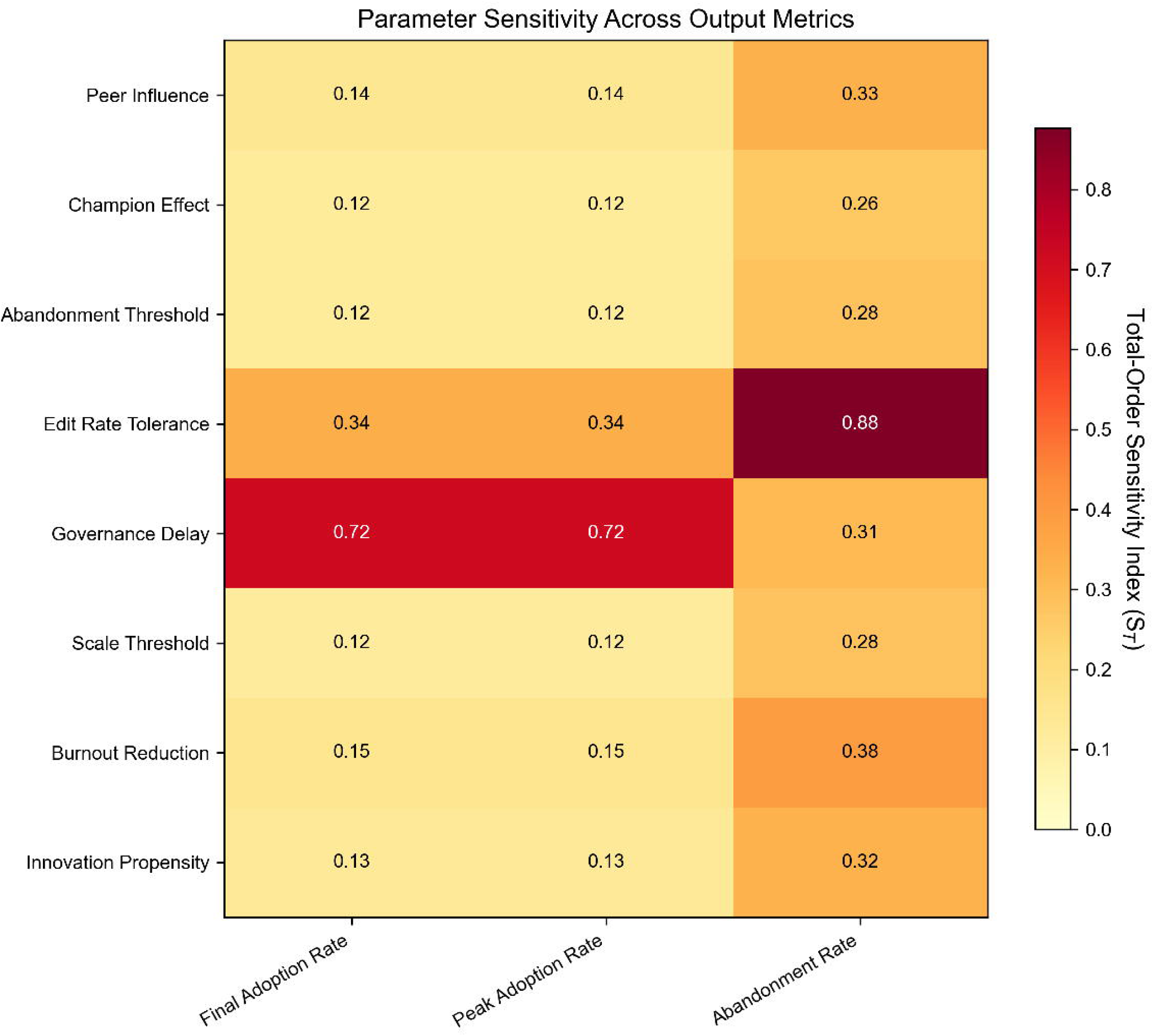
Parameter sensitivity heatmap showing total-order Sobol sensitivity indices (S_T_) for each parameter across multiple output metrics (final adoption rate, peak adoption rate, abandonment rate). Governance delay shows the largest effect on adoption outcomes; edit-rate tolerance dominates abandonment rate sensitivity. Champion rate indices were non-estimable due to near-zero variance and are omitted.

**Figure S4.**
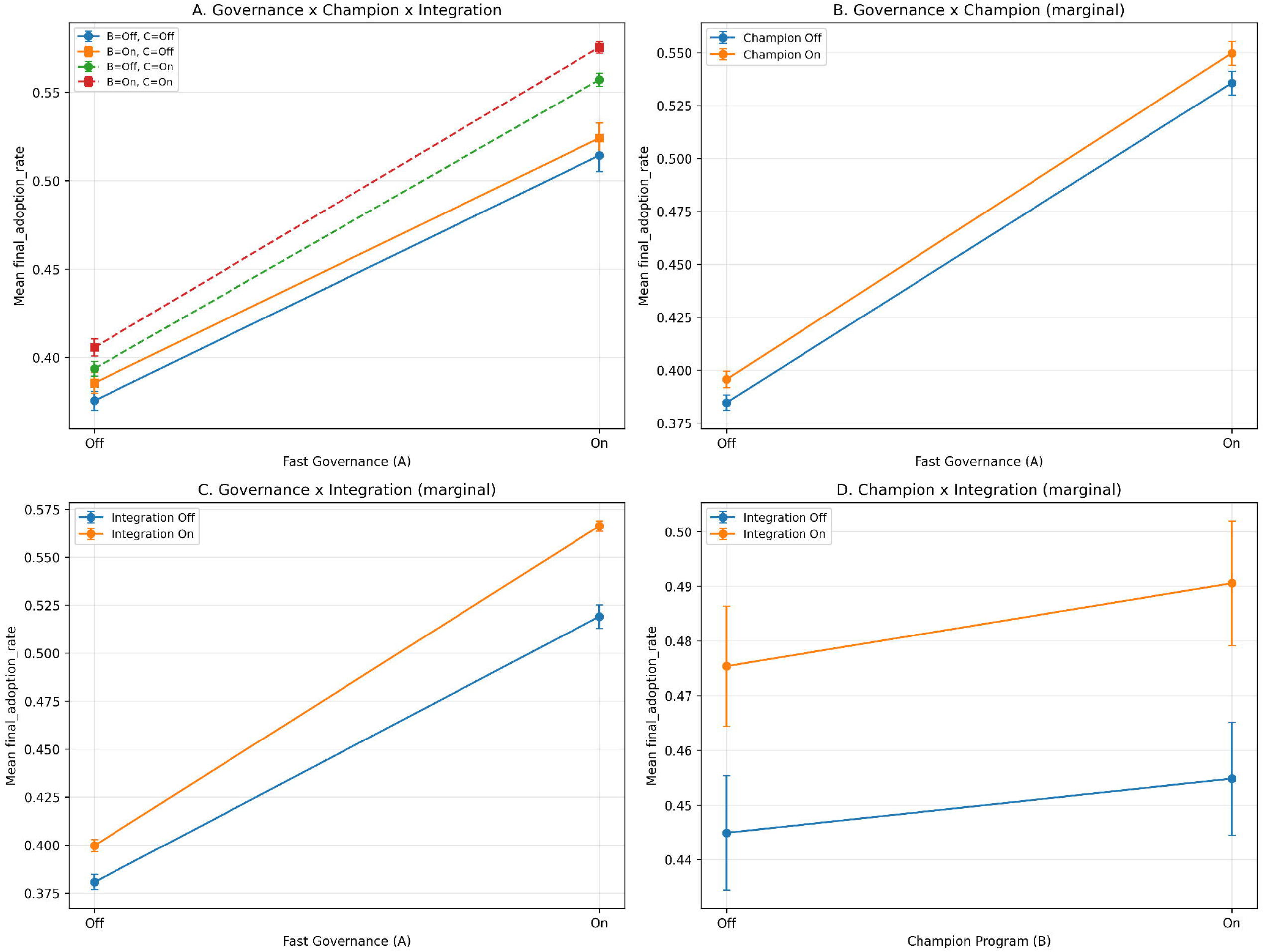
2^3^ Factorial interaction analysis. **(A)** Three-factor interaction plot showing mean final adoption rate by Fast Governance (A), Champion Program (B), and Better EHR Integration (C) condition. **(B)** Governance x Champion marginal interaction. **(C)** Governance x Integration marginal interaction. **(D)** Champion x Integration marginal interaction. Error bars represent standard errors of cell means across 30 realizations per condition. Non-parallel lines indicate interaction effects; see **Table S19** for formal ANOVA statistics.

