## Supplementary Methods for "An Agent-Based Modeling Framework for Healthcare AI Adoption: Application to Ambient Clinical Documentation"

### S1. Complete ODD Protocol

#### S1.1 Purpose

The purpose of this agent-based model is to understand the adoption dynamics of ambient clinical documentation AI tools in health systems. Specifically, the model aims to:

1. **Explain** how micro-level decisions by clinicians, organizations, and vendors aggregate to produce population-level adoption patterns
2. **Explore** how different policy interventions, technology improvements, and organizational strategies affect adoption dynamics and sustained use
3. **Predict** (qualitatively) adoption trajectories under different scenarios to inform decision-making by health system leaders

#### S1.2 Entities, State Variables, and Scales

##### Entities

| Entity | Description | Count (Baseline) | Count (Full Scale) |
| --- | --- | --- | --- |
| Clinician | Healthcare providers who may adopt the tool | 500 | 50,000 |
| Organization | Healthcare organizations that govern access | 10 | 500 |
| Vendor | Technology vendors offering ambient scribe products | 4 | 4 |
| Network | Peer influence network connecting clinicians | 1 | 1 |

##### Clinician State Variables

| Variable | Type | Description | Initial Value |
| --- | --- | --- | --- |
| adoption_state | Categorical | Current adoption status | never_tried |
| psychotype | Categorical | Innovation propensity category | Sampled from distribution |
| specialty | Categorical | Clinical specialty | Sampled from distribution |
| innovation_propensity | Float [0,1] | Individual tendency to try new technology | Beta(2.0, 5.0) |
| friction_tolerance | Float [0,1] | Tolerance for integration friction | Beta(2.0, 2.0) |
| benefit_threshold | Float [0,1] | Minimum perceived benefit required | Uniform(0.1, 0.5) |
| burnout_level | Float [0,1] | Current burnout level | Beta(3.0, 2.0) |
| cumulative_benefit | Float | Accumulated positive experience | 0.0 |
| cumulative_friction | Float | Accumulated negative experience | 0.0 |
| weeks_as_user | Integer | Duration in current adoption state | 0 |
| habit_strength | Float [0,1] | Usage habit strength | 0.0 |

**Adoption States:** - never_tried: Has not attempted tool use - aware_not_tried: Aware but has not initiated trial - trialing: Currently in trial period - high_user: Regular user (>78% encounter usage) - medium_user: Moderate user (30-78% encounter usage) - low_user: Low-intensity user (<30% encounter usage) - champion: Promoter after sustained positive experience - abandoned: Former user who discontinued - detractor: Active discourager after negative experience

**Psychotype Categories (adapted from Rogers 2003):** - high_need_early_adopter (15%): High documentation burden sensitivity, high risk tolerance, analogous to Rogers’ Innovators (expanded from 2.5% to 15% to reflect healthcare documentation urgency) - evidence_driven_pragmatist (40%): Evidence-driven, peer-influenced, analogous to Rogers’ Early Majority - compliance_oriented_late_adopter (30%): Compliance-focused, requires organizational sanction, analogous to Rogers’ Late Majority - resistant_non_adopter (15%): Low adoption probability regardless of evidence, analogous to Rogers’ Laggards

**Specialty Categories:** - primary_care (40%): Highest documentation burden, benefit multiplier 1.2 - specialty_medical (30%): Average burden, benefit multiplier 1.0 - surgical (20%): Lower documentation burden, benefit multiplier 0.8 - emergency (10%): Lowest documentation burden, benefit multiplier 0.6

##### Organization State Variables

| Variable | Type | Description | Initial Value |
| --- | --- | --- | --- |
| phase | Categorical | Deployment phase | not_started |
| integration_maturity | Float [0,1] | EHR integration infrastructure | Beta(3.0, 2.0) |
| governance_delay_weeks | Integer | Time for approval | Uniform(8, 28) |
| pilot_duration_weeks | Integer | Planned pilot length | Uniform(13, 52) |
| scale_threshold | Float [0,1] | Retention threshold for scaling | 0.50 |
| current_vendor_id | Integer | Selected vendor | None |

**Organization Phases:** - not_started: No evaluation initiated - shadow_it: Informal clinician use detected - evaluating: Vendor assessment in progress - security_review: Security review phase - governance: Awaiting approval (time-gated) - pilot: Limited deployment for evaluation - pilot_stagnation (pilot_purgatory in code): Modeled pilot stagnation state entered when pilot retention is 0.20-0.55; organizations may exit to scaling or termination via a retention- and time-dependent hazard - scaling: Expanding beyond pilot - full_deployment: Enterprise-wide availability - terminated: Deployment discontinued

##### Vendor State Variables

| Variable | Type | Description | Initial Value |
| --- | --- | --- | --- |
| performance_score | Float [0,1] | Product accuracy | Beta(3.0, 2.0) |
| integration_quality | Float [0,1] | EHR integration depth | Beta(4.0, 2.0) |
| base_edit_rate | Float [0,1] | Baseline note edit rate | Beta(2.5, 7.5) ~25% |
| annual_price | Float | Per-clinician annual cost | Uniform(9000, 11000) |

##### Scales

- **Temporal**: Discrete weekly time steps; default horizon 104 weeks (2 years)
- **Spatial**: Not explicitly spatial; network topology represents social connections
- **Population**: Scalable from 500 to 50,000+ clinicians

#### S1.3 Process Overview and Scheduling

Each simulation week proceeds in the following order:

1. **Organization decisions**: Phase transitions (evaluate → governance → pilot → scale)
2. **Vendor decisions**: Annual price and integration updates (every 52 weeks)
3. **Clinician decisions**:
   - Trial initiation (for never_tried)
   - Experience accumulation (for trialing/active users)
   - State transitions (adoption, partial use, abandonment)
   - Champion/detractor emergence
4. **State updates**: Increment time-in-state counters, update habit strength

#### S1.4 Design Concepts

##### Theoretical Background

The model integrates: - **Diffusion of innovations** (Rogers, 2003): Psychotype heterogeneity, adopter categories, S-curve dynamics - **Technology adoption in healthcare** (Greenhalgh et al., 2004): Organizational context, governance processes - **Peer influence in physician behavior** (Donohue et al., 2018): Network-mediated adoption spillovers

##### Individual Decision-Making

**Clinicians** make bounded rational decisions based on: - Personal characteristics (psychotype, friction tolerance, benefit threshold) - Peer influence (weighted observation of network neighbors) - Accumulated experience (benefit vs. friction balance) - Learning curve effects (friction and edit rates decline with time-in-use) - Access constraints (organizational phase, pilot membership) - Pilot enrollment effect (minimum weekly trial probability for pilot cohort clinicians)

**Organizations** make sequential decisions: - Vendor selection (price, integration quality, performance weighted) - Governance approval (time-delayed gating) - Scale decisions (pilot retention threshold)

##### Emergence

The following system-level outcomes emerge from agent interactions: - Adoption S-curves with characteristic inflection points - Organizational outcome heterogeneity (success, pilot stagnation, failure) - Champion clustering and cascade dynamics - Specialty-stratified adoption patterns

##### Stochasticity

Stochasticity enters through: 1. **Agent initialization**: Characteristics drawn from specified distributions 2. **Network construction**: Watts-Strogatz rewiring 3. **Decision rules**: Probabilistic trial initiation, experience noise 4. **Technical issues**: Random friction spikes

### S2. Parameter Tables with Empirical Sources

#### Table S1. Clinician Parameters

| Parameter | Value/Distribution | Empirical Source |
| --- | --- | --- |
| Psychotype: high_need_early_adopter | 15% | Rogers (2003), adapted |
| Psychotype: evidence_driven_pragmatist | 40% | Rogers (2003), adapted |
| Psychotype: compliance_oriented_late_adopter | 30% | Rogers (2003), adapted |
| Psychotype: resistant_non_adopter | 15% | Rogers (2003), adapted |
| innovation_propensity | Beta(2.0, 5.0) | Model calibration |
| friction_tolerance | Beta(2.0, 2.0) | Model calibration |
| benefit_threshold | Uniform(0.1, 0.5) | Model calibration |
| Initial burnout | Beta(3.0, 2.0), mean ≈ 0.6 | AMA surveys: 62.8% peak burnout |
| burnout_reduction_per_use | 0.005 | Target: 13.1pp reduction (Olson et al. 2025) |
| edit_rate_tolerance | Normal(0.45, 0.08) | Mayo Clinic: 42% threshold |
| trial_uses_to_decision | 10 encounters | Stanford qualitative study |
| weeks_to_champion | 12 weeks | Model assumption (3 months) |

#### Table S2. Organization Parameters

| Parameter | Value/Distribution | Empirical Source |
| --- | --- | --- |
| governance_delay_weeks | Uniform(8, 28) | KLAS: 6.6 months average |
| security_review_weeks | Uniform(12, 26) | Industry: 3-6 months |
| pilot_duration_weeks | Uniform(13, 52) | Literature: 3-12 months |
| pilot_target_size | 50 clinicians | Literature: 50 typical |
| scale_threshold | 0.50 | >50% retention for scaling |
| stagnation_exit_hazard | 0.02/week | Model assumption (pilot stagnation exit hazard) |
| stagnation_scale_probability | 0.30 | Model assumption (scale vs terminate) |
| stagnation_retention_floor | 0.20 | Model assumption (immediate terminate) |
| stagnation_retention_ceiling | 0.55 | Model assumption (immediate scale) |
| pilot_trial_min_probability | 0.50 | Pilot enrollment expectation (model assumption) |
| integration_maturity | Beta(3.0, 2.0) | Model calibration |

#### Table S3. Vendor Parameters

| Parameter | Value/Distribution | Empirical Source |
| --- | --- | --- |
| annual_price_enterprise | Uniform(9000, 11000) | Becker’s: $9,000+/year |
| monthly_price_individual | Uniform(120, 600) | Market: $120-600/month |
| base_edit_rate | Beta(2.5, 7.5), mean ≈ 0.25 | Literature: ~25% baseline |
| performance_score | Beta(3.0, 2.0) | Model calibration |
| integration_quality | Beta(4.0, 2.0) | Model calibration |

#### Table S4. Network and Interaction Parameters

| Parameter | Value | Empirical Source |
| --- | --- | --- |
| Network type | Small-world (Watts-Strogatz) | Barnett et al. (2011) |
| k_neighbors | 6 | Model calibration |
| rewiring_probability | 0.1 | Standard small-world |
| peer_influence_coefficient | 0.07 | Donohue et al. (2018): 5.9-8.3% |
| champion_influence_multiplier | 2.0 | Borracci & Giorgi (2018): ~1.2-1.5x; 2.0 used as optimistic proxy |
| detractor_influence_multiplier | 1.5 | Model assumption |

#### Table S5. Adoption Dynamics Parameters

| Parameter | Value | Empirical Source |
| --- | --- | --- |
| abandonment_friction_threshold | 0.82 | Model calibration |
| habit_growth_rate | 0.08/week | Model calibration |
| habit_cap | 0.70 | Model calibration |
| base_hazard_rate | 0.033/week | Calibration prior for post-adoption dropout |
| early_trial_weeks | 8 | Model assumption |
| high_use_threshold | 0.78 | Stanford: >78% encounters |
| medium_use_threshold | 0.30 | Stanford: 30-78% |

### S3. Calibration Targets

#### Table S6. Empirical Calibration Targets

| Target | Empirical Value | Source |
| --- | --- | --- |
| Clinician adoption (low support) | 20-40% | Published case studies and surveys (see main References 4-5) |
| Clinician adoption (standard) | 40-55% | Published case studies and surveys (see main References 4-5) |
| Clinician adoption (intensive) | 65-80% | Large enterprise deployments and surveys (see main References 2, 4) |
| Pilot-to-scale attrition | 35-45% | Intermountain cohort study (main Reference 5) |
| Usage: high users | ~41% | Stanford qualitative study (main Reference 13) |
| Usage: medium users | ~27% | Stanford qualitative study (main Reference 13) |
| Usage: low users | ~23% | Stanford qualitative study (main Reference 13) |
| Usage: dropout | ~9% | Stanford qualitative study (main Reference 13) |
| Time savings | ~5 min/encounter | Intermountain cohort and related reports (main Reference 5) |
| Burnout reduction | 13.1pp over 30 days | Olson et al. (main Reference 17) |

*Note: These targets are used as calibration priors and include values drawn from peer-reviewed studies and industry reports; the latter may not have DOIs.*

S4. Sensitivity Analysis Methodology

#### S4.1 Parameter Selection

Eight parameters were selected for sensitivity analysis based on preliminary screening and theoretical importance:

1. **abandonment_friction_threshold**: Friction level triggering user abandonment
2. **governance_delay_mean**: Average weeks for organizational approval
3. **edit_rate_tolerance_mean**: Average acceptable edit rate before abandonment
4. **peer_influence_coefficient**: Strength of peer adoption effect
5. **innovation_propensity_alpha**: Shape parameter for innovation propensity distribution
6. **scale_threshold**: Pilot retention threshold for organization scaling
7. **burnout_reduction_per_use**: Per-use reduction in clinician burnout
8. **champion_influence_multiplier**: Influence amplification for champion clinicians

#### S4.2 Sampling Design

We used Saltelli’s extension of Sobol’ sequences for efficient variance-based sensitivity analysis: - Base samples: 64 - Total parameter sets: N(2k+2) = 64(2×8+2) = 1,152 - Replicates per parameter set: 2 (2,304 total simulations) - Output metrics: final_adoption_rate and abandonment_rate reported; peak_adoption_rate and champion_rate were explored but not emphasized due to instability or non-estimable variance

#### S4.3 Index Computation

**First-order index (S1)**: Variance contribution from parameter alone

$$S_{i}=\frac{V_{X_{i}}(E_{X_{\sim i}}(Y|X_{i}))}{V(Y)}$$

**Total-effect index (ST)**: Variance contribution including all interactions

$$ST_{i}=\frac{E_{X_{\sim i}}(V_{X_{i}}(Y|X_{\sim i}))}{V(Y)}$$

#### S4.4 Confidence Intervals

Bootstrap confidence intervals (100 resamples; SALib default) were computed for all indices.

### S5. Scenario Specifications

#### Table S7. Intervention Scenario Definitions

| Scenario | Parameter Changes | Rationale |
| --- | --- | --- |
| **Fast-Track Governance** | governance_delay_weeks: Uniform(8,28) → Fixed(4) | Streamlined approval process |
| **Champion Program** | peer_influence_coefficient: 0.07 → 0.12 | Formal peer support program |
| **Better EHR Integration** | base_edit_rate: Beta(2.5,7.5) → Fixed(0.15) | Improved vendor integration |
| **Reduced Resistance (proxy)** | resistant_non_adopter_fraction: 0.15 → 0.0; evidence_driven_pragmatist: 0.40 → 0.55 | Proxy for lowering clinician resistance (not a coercive mandate) |
| **High Burnout Crisis** | burnout_distribution: Beta(3,2) → Normal(0.70, 0.10) | Elevated baseline burnout |
| **Combined Intervention** | Fast governance + Champion + Better integration | Multi-lever intervention |

#### Table S8. Intervention Magnitudes Relative to Baseline

| Scenario | Baseline value | Intervention value | Percent change |
| --- | --- | --- | --- |
| **Fast-Track Governance** | 18 weeks (mean) | 4 weeks | -78% |
| **Champion Program** | 0.07 | 0.12 | +71% |
| **Better EHR Integration** | 0.25 edit rate (mean) | 0.15 | -40% |
| **Reduced Resistance (proxy)** | 15% resistant non-adopters | 0% | -100% |
| **High Burnout Crisis** | 0.60 mean burnout | 0.70 | +17% |

*Note: Percent change is relative to baseline mean values; the Combined Intervention bundles multiple changes.*

### S6. Pattern Plausibility Approach

#### S6.1 Pattern-Oriented Modeling (POM)

We employed pattern-oriented modeling to validate against multiple empirical patterns simultaneously:

**Micro-level patterns:** - Psychotype ordering: High-need early adopters adopt before evidence-driven pragmatists before compliance-oriented late adopters before resistant non-adopters

**Meso-level patterns:** - Pilot-to-scale attrition: 35-45% of pilot users should discontinue or underutilize (completed trialers; ongoing trialing excluded)

**Macro-level patterns:** - Adoption curve shape: S-curve with characteristic inflection - Specialty ordering: Primary care > specialty medical > surgical > emergency - Usage distribution: Trimodal distribution matching Stanford qualitative findings

These patterns were prespecified and evaluated on a separate run set; because some overlap with calibration priors, they are interpreted as directional consistency checks rather than independent validation.

#### S6.2 Plausibility Check Results

| Pattern | Expected | Observed | Status |
| --- | --- | --- | --- |
| Psychotype ordering | Early Adopter > Pragmatist > Late Adopter > Non-Adopter | Correct ordering (ρ=1.0) | PASS |
| Pilot attrition (completed trialers) | 35-45% | 35.5% | PASS |
| Adoption inflection | Week 52-104 | Week 91.3 (mean) | PASS |
| Specialty ordering | PC > SM > Surg > EM | ρ=1.0 | PASS |
| Usage distribution (completed trialers) | High 35-50%, Medium 20-35%, Low 15-30%, Dropout 5-15% | High 35.2%, Medium 29.5%, Low 23.1%, Abandoned 12.1% | PASS |
| Mature user dropout | 5-15% | 7.7% | PASS |

All six plausibility patterns passed their prespecified criteria.

Validation simulations were generated using scripts/tiered_validation.py with configs/calibrated_set_f.yaml (50,000 clinicians, 500 organizations; seeds 42-71).

### S7. Computational Details

#### S7.1 Implementation

- **Language**: Python (reported runs executed in Python 3.12)
- **Dependencies**: NumPy 1.26+, NetworkX 3.2+, Pydantic 2.5+, Pandas 2.1+, Matplotlib 3.8+, SALib 1.4+
- **Random number generation**: NumPy Generator with explicit seeding

#### S7.2 Computational Resources

- **Hardware**: Apple M-series or equivalent
- **Runtime**: ~4 minutes per realization (50,000 clinicians, 104 weeks; 4 workers)
- **Sensitivity analysis**: ~35 hours (2,304 simulations across 1,152 parameter sets, 4 parallel workers)
- **Emergent analysis**: ~29 minutes (30 realizations, 4 parallel workers)

#### S7.3 Reproducibility

All analyses use explicit random seeds. Configuration files and analysis scripts are provided in the code repository. Stored run manifests capture Python version, platform, and package versions; git commit hashes were not recorded for the reported runs.

### S8. Quantitative Validation Methodology

#### S8.1 Rationale

To address the absence of direct empirical validation, we compared the model’s simulated output distributions against published deployment data from ambient clinical documentation implementations at seven health systems and three aggregate industry benchmarks. This comparison is not calibration in the parameter-fitting sense: mechanistic parameters (peer influence coefficients, edit-rate thresholds, psychotype distributions) were derived from literature and were not tuned to match these deployment outcomes. The one exception is the optional staggered-initialization regime, whose starting-phase distribution is calibrated to verified governance-to-deployment timelines (see below); this targets initial conditions, not the model’s behavioral parameters. The comparison assesses whether published real-world values fall within the model’s predictive distribution and quantifies the discrepancy where they do not.

All cited deployment values were independently re-verified against primary sources prior to this analysis. The verification report (docs/empirical_verification_report.md in the public release) records the source, exact figures, and verification status for each value. Three changes resulted: (i) the Ambience ~50% retention figure, a non-peer-reviewed industry claim, was removed as a comparison anchor; (ii) the UCSF “20-week learning period” attribution was corrected (the cited Holmgren et al. study reports physician financial-productivity outcomes, not a learning-period metric); and (iii) the Cleveland Clinic 80% value was downgraded to an illustrative exemplar because it could not be traced to a named primary source and may coincide with an anonymized industry benchmark.

#### S8.2 Reference-Class Separation

The model simulates an ecosystem of 500 heterogeneous organizations with varying governance timelines, vendor selections, and implementation intensities. Published deployment data fall into three distinct reference classes that correspond to different model quantities and must not be pooled: (a) early single-site trajectories (adoption measured over time at one motivated site, e.g., VUMC, Kaiser), compared against the modeled ecosystem trajectory; (b) mature single-site final adoption (e.g., VUMC 51%, Kaiser 73%, Atrium 75%), an upper-tail reference; and (c) cross-sectional or randomized ecosystem snapshots (UCLA RCT 32%, PHTI 20-50%, Menlo 35%), the closest analogue to the modeled ecosystem mean. We report goodness-of-fit separately for each class.

#### S8.2a Initialization Regimes

We evaluate two initialization regimes. In the **cold-start** regime (default), all organizations begin in a not-started state. In the **staggered** regime, organizations are seeded at t=0 into a distribution of deployment phases, with clinician adoption states for past-governance organizations drawn from a phase-consistent distribution (active-use fractions split high/medium/low per the Stanford pattern). The starting-phase distribution is calibrated to the verified governance-to-deployment timelines (Cleveland ~6 months, Vanderbilt ~10 months, Kaiser ~12 months, Mass General Brigham ~24 months), reflecting that most published systems were already mid-rollout when observed. This is implemented as a configuration flag (staggered_initialization) and unit-tested; the default behavior is unchanged.

#### S8.2b Plateau Re-calibration (Set G)

Staggered initialization removed the cold-start artifact but raised the steady-state plateau above the cross-sectional and randomized ecosystem benchmarks (final median ~72%). To align the early-time trajectory and the steady-state plateau with published data simultaneously, we performed a formal calibration over six free parameters:

| Parameter | Role | Schema bound | Calibrated value |
| --- | --- | --- | --- |
| base_hazard_rate | weekly post-adoption abandonment hazard | ≤ 0.05 | 0.0401 |
| abandonment_friction_threshold | friction level triggering abandonment | [0, 1] | 0.3949 |
| scale_threshold | org retention needed to scale from pilot | [0, 1] | 0.6197 |
| deployed_weight | mass on seeded deployed phases at init | derived distribution | 0.4309 |
| seeded_active_fraction_scaling | active-user fraction, SCALING orgs at t=0 | [0, 1] | 0.1412 |
| seeded_active_fraction_full | active-user fraction, FULL_DEPLOYMENT orgs at t=0 | [0, 1] | 0.5690 |

All other structural parameters (peer influence, champion multiplier, edit-rate tolerance distribution, psychotype proportions, governance-delay distributions) were held at their prior literature-anchored values. The calibration target was the cross-sectional ecosystem plateau (center 0.34, the midpoint of the UCLA randomized 32%, Menlo 35%, and PHTI 20-50% anchors), subject to two constraints: a rising early-time trajectory (not the near-zero cold-start artifact), operationalized as a lower bound on ecosystem adoption at week 8; and an organizational upper tail reaching the motivated single-site range (organization-level 90th-percentile final adoption above 0.50). We minimized a reference-class-weighted loss combining squared plateau error, an early-time shape penalty, and an upper-tail penalty.

The search used Latin-hypercube sampling: 100 coarse configurations (4 replicates each, 4,000 clinicians / 80 organizations), then 30 refined configurations (8 replicates, 8,000 clinicians / 160 organizations). An initial optimum required a base_hazard_rate of 0.12, which violates the configuration schema’s 5%-per-week cap on the weekly abandonment hazard; the search was re-run within schema-valid bounds, and we confirmed that the friction threshold and deployed-mass parameters – not the abandonment hazard – are the primary plateau levers, so the calibration target is reachable within valid bounds. The search established that the achievable ecosystem-mean plateau floor within valid bounds is approximately 0.40, but that the loss-optimal configuration reaches a median final adoption of 0.34 with a valid rising trajectory. The calibrated configuration (“set G”, configs/calibrated_set_g_recalibrated.yaml) was confirmed at full scale (50,000 clinicians, 500 organizations, 10 realizations: final median 0.364, mean 0.336, SD 0.114; 8/10 realizations inside the empirical cross-sectional band). The calibration harness (scripts/calibration_harness.py), search driver (scripts/run_calibration_search.py), and search results (results/calibration/) are included in the public release.

#### S8.2c Native Re-run of Interventions and Sobol Screen Under Set G

To make the results section natively calibrated rather than relying on the original configuration, we re-ran both the full six-scenario intervention comparison and the eight-parameter Sobol screen under set G (scripts recalibrated_interventions_full.py and sensitivity_setg.py; results in results/validation_quant/ and results/sensitivity_setg/).

The intervention re-run used paired contrasts (30 realizations per scenario, shared seeds with the baseline) at 8,000 clinicians / 160 organizations, the scale whose baseline adoption (34.0%) most closely matches the confirmed full-scale baseline (33.6%). We also ran the same six scenarios at 4,000/80 to characterize scale-dependence. The intervention effects are strongly scale-dependent: at 4,000/80 (baseline 31.8%, an outlier relative to full scale) the combined intervention gave +13.5pp and was clearly largest, whereas at the matched 8,000/160 scale all effects roughly halved and the combined intervention (+3.2pp) fell below fast-track governance alone (+4.6pp). Two independent runs (20 and 30 realizations) reproduced the combined ≤ fast-governance ordering, so it is not a sampling artifact. Because the manuscript scale is 500 organizations, the smaller-scale figures overstate intervention leverage; we therefore report the 8,000/160 results (main **Table 5**) as primary. At that scale only the reduced-resistance lever has a paired-d_z_ confidence interval excluding zero (d_z_=0.90 [0.57, 1.45]), because it acts uniformly on every clinician rather than through high-variance organizational gates; the other single-lever effects are directionally positive but not statistically distinguishable from zero.

The Sobol screen used the identical eight-parameter Saltelli design (64 base samples, 1,152 parameter sets, 2 replicates each) with the base configuration seeded from set G. The one adjustment was widening the abandonment_friction_threshold sampling box to [0.35, 0.90] so the calibrated operating point (0.395) falls inside the screened range. Total-order indices for final adoption are reported in **Table S21**. The dominant driver changes from governance delay in the original configuration (S_T_=0.72) to the abandonment/friction threshold under set G (S_T_=0.79), with edit-rate tolerance again in the top tier (S_T_=0.51) and governance delay third (S_T_=0.40). This reordering is mechanistically consistent with the calibration, which made clinician friction and churn the primary determinants of the steady-state plateau; edit-rate tolerance remains a top-tier driver across both configurations.

#### Table S21. Total-Order Sobol Indices for Final Adoption: Original vs. Re-calibrated

| Parameter | S_T_ (re-calibrated) | S_1_ (re-calibrated) | S_T_ (original) | S_1_ (original) |
| --- | --- | --- | --- | --- |
| Abandonment/friction threshold | 0.79 | 0.53 | 0.12 | 0.13 |
| Edit-rate tolerance (mean) | 0.51 | 0.24 | 0.34 | 0.26 |
| Governance delay (mean) | 0.40 | -0.02 | 0.72 | 0.57 |
| Peer influence coefficient | 0.40 | 0.12 | 0.14 | -0.01 |
| Champion influence multiplier | 0.37 | -0.12 | 0.12 | -0.02 |
| Pilot-to-scale threshold | 0.33 | 0.03 | 0.12 | -0.05 |
| Burnout reduction per use | 0.29 | -0.11 | 0.15 | 0.09 |
| Innovation propensity (alpha) | 0.23 | -0.07 | 0.13 | -0.03 |

Confidence intervals at N=64 base samples are wide (small negative S_1_ estimates are sampling noise around zero), so these indices should be read as an ordinal reordering of the leading drivers rather than precise magnitudes.

#### S8.3 Data Sources

| Site | Citation | N Clinicians | Observation Period | Key Metrics |
| --- | --- | --- | --- | --- |
| VUMC | Wright et al., JAMIA 2025 | 2,400 | 10.5 weeks | 9.7% day 1, 42% wk 6, 51% wk 10.5 |
| Kaiser/TPMG | Tierney et al., NEJM Catalyst 2025 | ~10,000 enabled (7,260 active) | 14 months | 72.6% final; MH>PC>EM specialty ordering |
| MGB | Mass General Brigham press releases (2023-2025) | 4,000 | ~28 months | 18→800→4,000 scale-up |
| Atrium Health | Liu et al., NEJM AI 2024 | 112 | Pilot | 75% active (>=25% notes), 60% high-use (>=60% notes) |
| Cleveland Clinic | Illustrative exemplar (unattributed; see S8.1) | N/R | Enterprise | ~80% utilization |
| UCLA | Lukac et al., NEJM AI 2025 (NCT06792890) | 238 | RCT | 30-34% utilization (non-self-selected) |
| Stanford | Shah et al., JAMA Network Open 2025 | 22 | Qualitative | 41/27/23/9% usage distribution |
| UCSF | Holmgren et al., JAMA Network Open 2026 | 1,565 | – | 44.6% physician adoption (productivity study; not a learning-period source) |
| PHTI | PHTI/Peterson Health Technology Institute, 2025 | N/A | Survey | 20-50% adoption range |
| Menlo Ventures | Survey (grey literature) | N/A | Survey | 35% average; 35-40% sustained |
| Ambience | *Removed* – unverified industry claim (was Fierce Healthcare 2025) | N/A | – | (excluded as anchor) |

#### S8.4 Comparison Dimensions

Eight dimensions were compared:

1. **Adoption at ~10 weeks**: Simulated adoption rate at week 10 vs. VUMC (51% at 10.5 weeks) and Kaiser (~34.4% extrapolated at ~10 weeks)
2. **Adoption at ~52 weeks**: Simulated adoption rate at week 52 vs. Kaiser (~60% at 12 months)
3. **Final adoption (104 weeks)**: Simulated final adoption rate vs. Kaiser (72.6%), VUMC (51%), Cleveland (80%), Atrium (75%), UCLA (32%), PHTI (20-50%), Menlo (35%)
4. **Usage distribution**: Simulated high/medium/low/dropout distribution vs. Stanford (41/27/23/9%) and Atrium (75% active, 60% high-use)
5. **Specialty ordering**: Simulated specialty adoption rates vs. Kaiser (mental health highest, with primary care and emergency both high) and literature consensus (PC>SM>Surg>EM)
6. **Governance timelines**: Simulated per-organization evaluation-to-pilot timeline vs. Cleveland (~6mo), VUMC (~10mo), Kaiser (~12mo), MGB (~24mo)
7. **Retention/plateau**: Simulated sustained use rate vs. PHTI (20-50%), Stanford/Atrium usage distributions, and Menlo (35-40%). (The Ambience ~50% retention figure used in the original submission was removed as an unverified industry claim; the retention dimension is now carried by the peer-reviewed usage-distribution anchors.)
8. **Curve shape**: Simulated inflection point timing vs. VUMC (S-curve) and Kaiser (phased). (The UCSF “20-week learning period” reference was removed; see S8.1.)

#### S8.5 Goodness-of-Fit Metrics

We replaced the descriptive range-overlap assessment used in the original submission with formal goodness-of-fit metrics. For each empirical observation we drew the model’s predictive distribution from 50 independent realizations per configuration (three configurations: cold-start configs/calibrated_set_f.yaml; staggered configs/calibrated_set_f_staggered.yaml; re-calibrated configs/calibrated_set_g_recalibrated.yaml), and computed:

- **RMSE / MAE**: root-mean-square and mean absolute error of the model’s predictive median against the observed value, aggregated within each reference class (adoption-rate units, 0-1).
- **90% predictive-interval coverage**: whether the observation falls within the model’s [5th, 95th] percentile band; reported as the fraction of observations covered per class. A well-calibrated nominal-90% interval covers ~0.9 of observations.
- **Posterior-predictive tail probability**: the two-sided tail probability of each observation under the model’s predictive distribution (2 x min(Pr[sim >= obs], Pr[sim < obs])); values near 0 indicate the observation lies in a distribution tail, values near 1 indicate central agreement.

Metrics were computed for all three configurations (cold-start, staggered, re-calibrated) so that the contribution of each successive correction is explicit, with 1,000-resample bootstrap confidence intervals on each per-class RMSE. The hardened validation script (scripts/hardened_validation.py), the earlier two-regime script (scripts/quantitative_validation.py), and the per-observation results (results/validation_quant/) are included in the public release. Headline results appear in main-text **Table 3** and **Figure 4**.

These analyses were run at a reduced scale (4,000-8,000 clinicians, 80-160 organizations) that reproduces the full-scale baseline final adoption (~38% vs. 37.6% at 50,000 clinicians) while making the three-configuration, 50-realization comparison computationally tractable; the re-calibrated configuration was additionally confirmed at full scale (S8.2b). We note that on the properly reference-class-matched anchors (cross-sectional ecosystem plus mature single-site; seven anchors), the re-calibrated model’s 90% predictive intervals cover 6/7 (86%), close to the nominal 90%, versus 2/7 for cold-start and 0/7 for staggered-only.

#### S8.6 Limitations of Retrospective Comparison

1. **Post-hoc design**: The comparison was not preregistered; dimensions and sites were selected after model development
2. **Selection bias**: Published deployment data overrepresent successful, well-resourced implementations
3. **Scale mismatch**: Model ecosystem (500 orgs) vs. single-site trajectories
4. **Implementation heterogeneity**: Sites differ in vendor, rollout strategy, support intensity, and mandated vs. voluntary adoption
5. **Temporal coverage**: Most published data cover <15 months; the model simulates 24 months
6. **Partial calibration**: Six free parameters were calibrated to the adoption-level and trajectory-shape reference classes (S8.2b); the resulting fit metrics for those classes therefore reflect in-sample calibration, not out-of-sample validation. The early single-site trajectory class was not a calibration target and functions as a held-out reference (all configurations miss it at the ecosystem level; only the re-calibrated model covers the corresponding organization-level quantity). Structural parameters (peer influence, psychotypes, governance-delay distributions, network topology) were not calibrated to these comparisons. The intervention comparison and Sobol screen were re-run natively under the calibrated configuration (S8.2c); the 2^3^ factorial analysis (Table S19) was retained from the original configuration and its interaction estimates were not re-derived under re-calibration.

#### S8.7 KLAS Extracted Values Note

Key values extracted from subscription-access industry reports: KLAS (2024) reported a mean healthcare AI procurement timeline of ~6.6 months across surveyed organizations; KLAS (2025) reported specialty-level adoption ordering with primary care leading. Exact extracted values are documented in parameter Tables S1-S4 to support reproducibility without requiring subscription access.

### S9. Factorial Interaction Analysis

#### S9.1 Methodology

To formally test for interactions among the three intervention factors used in the combined scenario, we conducted a 2^3^ full factorial analysis crossing:

- **Factor A: Fast Governance** — governance_delay reduced from baseline Uniform(8, 28) weeks to fixed 4 weeks
- **Factor B: Champion Program** — peer_influence_coefficient increased from 0.07 to 0.12
- **Factor C: Better EHR Integration** — vendor base_edit_rate reduced from baseline Beta(2.5, 7.5) (~0.25 mean) to fixed 0.15

This produced 8 conditions (all combinations of factor on/off). Each condition was run with 30 realizations using matched random seeds (base seed = 42), yielding 240 total simulations. The primary outcome was final adoption rate at week 104.

#### S9.2 Analysis

Main effects, two-way interactions, and the three-way interaction were estimated via factorial ANOVA with Type III sums of squares. Effect sizes are reported as partial eta-squared (SS_effect / (SS_effect + SS_residual)). Interaction significance was assessed using F-tests with alpha = 0.05.

#### Table S19. 2^3^ Factorial ANOVA: Final Adoption Rate

| Source | SS | df | MS | F | p | Partial η² |  |  |
| --- | --- | --- | --- | --- | --- | --- | --- | --- |
| A (Fast Governance) | 1.395 | 1 | | 1.395 | 1317.98 | | <0.001 | 0.850 |
| B (Champion Program) | 0.009 | 1 | | 0.009 | 8.91 | | 0.003 | 0.037 |
| C (Better EHR Integration) | 0.066 | 1 | | 0.066 | 62.06 | | <0.001 | 0.211 |
| A × B | 0.000 | 1 | | 0.000 | 0.14 | | 0.713 | 0.001 |
| A × C | 0.012 | 1 | | 0.012 | 11.13 | | 0.001 | 0.046 |
| B × C | 0.000 | 1 | | 0.000 | 0.40 | | 0.530 | 0.002 |
| A × B × C | 0.000 | 1 | | 0.000 | 0.16 | | 0.690 | 0.001 |
| Residual | 0.246 | 232 | | 0.001 | — | | — | — |

*Note: Type III sums of squares. 30 realizations per condition, matched seeds (base_seed=42). Fast governance dominates (partial η²=0.850); the only significant interaction is governance × integration (A × C, partial η²=0.046). See Figure S4 for interaction plots.*

#### S9.3 Interpretation

The factorial design replaces the informal comparison of combined-vs-sum-of-individuals used in the initial submission with proper statistical tests for interaction effects. Fast governance was the dominant factor (partial η²=0.850), accounting for the majority of variance in final adoption rate. The only significant interaction was governance × EHR integration (A × C; F(1,232)=11.13, P=0.001, partial η²=0.046): fast governance amplified the benefit of lower edit rates (effect of governance when integration is on: +16.7pp vs. off: +13.9pp). The three-way interaction was not significant (F=0.16, P=0.69), indicating that the combined intervention’s advantage reflects additive main effects plus this specific pairwise synergy rather than emergent three-way dynamics. Cell means ranged from 0.376 (baseline) to 0.575 (all three interventions active).

#### Table S20. Calibration–Plausibility Mapping

| Data Source | Used for | Held out from |
| --- | --- | --- |
| Rogers diffusion proportions (2003) | Calibration (psychotype fractions) | — |
| Peer influence 5.9-8.3% (Donohue 2018) | Calibration (peer coefficient) | — |
| Pilot-to-scale attrition 35-45% (Haberle 2024) | Calibration target | — |
| Usage distribution 41/27/23/9% (Shah 2025) | Calibration target | — |
| KLAS governance ~6.6 mo (2024) | Calibration (governance delay) | — |
| Edit-rate tolerance ~40% (Shah 2025) | Calibration (edit threshold) | — |
| Specialty ordering PC>EM (KLAS 2025) | Plausibility check pattern | Calibration |
| Psychotype adoption ordering | Plausibility check pattern | Calibration |
| Adoption inflection ~15 mo (Tierney 2025) | Plausibility check pattern (directional) | Calibration |
| Published deployment data (7 sites + 3 benchmarks) | Retrospective plausibility check | Calibration + plausibility checks |

*Note: This table maps each empirical data source to its role in model development. Sources used for calibration were not independently used as plausibility checks. The “held out from” column indicates which model development stages did not use that source.*

### S10. Extended Discussion

#### S10.1 ABM Methodology Comparison

Agent-based modeling offers advantages over alternative approaches for this problem domain. Traditional diffusion models (Bass, logistic growth) assume population homogeneity and do not capture the psychotype-specific adoption dynamics observed here. System dynamics models aggregate individual decisions into stock-flow structures, which can obscure the network effects and emergent organizational outcomes that characterize healthcare AI adoption (main Reference 22). Our approach aligns with calls for complexity-aware methods in health technology assessment. Greenhalgh and colleagues’ NASSS framework emphasizes that technology adoption involves interactions between adopters, organizations, the wider system, and the technology itself (main Reference 23). These are multi-level dynamics well suited to agent-based modeling. The model’s reproduction of empirical patterns (psychotype ordering, specialty variation, S-curve dynamics) alongside novel predictions (organizational outcome distributions, intervention interactions) illustrates the potential value of simulation-based approaches for scenario exploration and hypothesis generation.

#### S10.2 Expanded Plausibility Interpretation

Pattern plausibility checks were prespecified from empirical reports and evaluated on a separate run set (main text Table 2; Supplementary Methods S6.2). The model reproduces ordering patterns for both psychotype and specialty and approximates usage distribution benchmarks, with pilot attrition within the prespecified range and inflection timing varying widely across run sets. The retrospective comparison against published deployment data from seven health systems (main text Table 3, Figure 4) provides additional context: the model’s ecosystem-level outputs are consistent with aggregate industry benchmarks (PHTI 20-50%, Menlo 35%) and non-self-selected populations (UCLA RCT 32%), while diverging from single-site champion implementations (Kaiser 73%, Cleveland Clinic 80%) in ways that reflect the model’s inclusion of organizational heterogeneity and governance delays.

Several important limitations apply to this comparison. First, it is post-hoc: the comparison was designed after both the model and the published data existed, precluding preregistered predictions. Second, published deployment data reflect selection bias toward successful implementations; health systems that abandoned ambient scribe deployments are underrepresented in the literature. Third, site-specific factors (vendor choice, implementation intensity, organizational culture, patient population) are not individually modeled; the comparison assesses whether published outcomes are plausible within the model’s ecosystem-level distribution rather than whether the model can reproduce any specific site’s trajectory. Fourth, the model represents a voluntary, phased adoption process, whereas some published sites used phased mandates or structured rollout schedules.

The model’s ecosystem-level adoption rate (simulated median 36.0% at 104 weeks) was lower than site-specific final rates reported by VUMC (51%), Kaiser (72.6%), Cleveland Clinic (80%), and Atrium (75%). This divergence is expected and informative: published reports describe well-resourced early adopters with dedicated implementation support, whereas the model’s ecosystem includes organizations that never progress past governance (the 49% pilot stagnation finding (rounded)). The UCLA RCT rate (32%), representing non-self-selected clinicians, the PHTI industry benchmark (20-50%), and the Menlo survey average (35%) all fell within or near the simulated range. Early adoption (weeks 10 and 52) was substantially lower in the simulated ecosystem than in published single-site data, reflecting the model’s inclusion of organizations still navigating governance and security review processes at those time points.

Usage intensity distributions approximated the Stanford four-category pattern: simulated high-use 35.2% vs. Stanford 41%; simulated medium-use 29.5% vs. Stanford 27%; simulated low-use 23.1% vs. Stanford 23%; simulated dropout 12.1% vs. Stanford 9% (plausibility check run set; Table 2, Table S12). Specialty ordering reproduced the expected documentation-burden gradient (Spearman rho = 1.0). Governance timelines (simulated median 11.1 months) overlapped with VUMC (10 months) and Kaiser (12 months). Retention/sustained use (simulated median 50.5%) overlapped with VUMC (51%) and Menlo (35-40%) benchmarks.

#### S10.3 Expanded Limitations

**Model validation.** The model should be interpreted as a theoretical framework for exploring adoption dynamics rather than a calibrated forecasting tool. We did not report adoption conditional on governance approval; future analyses could isolate post-gate dynamics as a negative-control framing. While parameters were grounded in empirical estimates where available, many required calibration assumptions. Screening-level Sobol analysis suggests governance delay and edit-rate tolerance are influential, but confidence intervals are wide, some first-order indices are negative, and rankings are not statistically distinguishable with N=64 base samples. Additional sampling would reduce uncertainty.

**Detractor dynamics.** No detractors emerged in the current run set, likely reflecting a modeling choice that routes dissatisfied users to abandonment rather than persistent vocal criticism. Although the model includes detractor transition logic (triggered by sustained high friction or edit rates exceeding individual tolerance), baseline vendor edit rates (~25%) rarely exceed clinician tolerance thresholds (~42%), making detractor transitions rare. This likely inflates net peer influence by eliminating negative word-of-mouth. A scenario with higher baseline edit rates or explicit detractor seeding would test the sensitivity of adoption outcomes to negative peer effects; this remains future work. Combined with the optimistic champion multiplier, this likely inflates peer-support effects and limits the model’s ability to capture negative word-of-mouth cascades.

**Initialization assumptions.** The model initializes all organizations in a not-started state, which prevents early-time adoption comparison with published sites that already had pilots underway. Future iterations could initialize a fraction of organizations at later deployment phases to enable early-trajectory comparison.

**Evidence heterogeneity.** Several calibration targets draw on industry reports or proxy measures (e.g., prescribing network peer effects), which may not transfer directly to ambient documentation adoption. The champion influence multiplier is an optimistic stress-test (2.0) relative to empirical ranges in prior diffusion models (approximately 1.2-1.5x; see Supplementary Table S4); sensitivity analysis suggests a modest total-effect index (S_T_=0.12), but the assumption remains a source of uncertainty. Peer influence estimates may overstate effects in high-friction technology adoption. These inputs should be validated against ambient-specific longitudinal data as it becomes available.

**Network structure.** The small-world network topology, while consistent with physician network studies (main Reference 15), represents a simplification of actual professional relationships. Network topology parameters (k=6 neighbors, rewiring probability 0.1) were not varied in sensitivity analysis. Because peer influence is a key adoption driver, adoption dynamics may differ under alternative network structures (e.g., scale-free or clustered networks). The model supports alternative topologies, but systematic topology sensitivity analysis remains future work. Organizational boundaries dominate tie formation in the model, with limited cross-organization ties. Barnett and colleagues found that shared patient relationships create stronger ties than geographic proximity; multi-layer networks incorporating referral patterns, training cohorts, and organizational hierarchies might capture additional peer influence pathways.

**Scope limitations.** The model focuses on clinician adoption decisions and does not explicitly represent patient outcomes, quality metrics, or cost-effectiveness, factors that may influence organizational decisions. Palm and colleagues’ finding that hallucinations were detected in 31% of ambient notes versus 20% of physician-authored notes (main Reference 21) raises safety considerations not addressed here. The model treats ambient documentation as a homogeneous technology class, whereas vendor-specific differences likely contribute to adoption heterogeneity. Application to other healthcare AI technologies (diagnostic AI, clinical decision support, administrative automation) would require domain-specific parameter estimation.

#### S10.4 Extended Model Insights

**Organizational heterogeneity.** Prior studies have documented individual clinician adoption variability (20-74%), but the organizational-level distribution, with success, pilot stagnation, and failure as distinct attractor states, represents a model prediction. The plurality of pilot stagnation outcomes (48.9%) suggests that organization-level factors may explain as much variance as individual characteristics. While governance delays are acknowledged in implementation reports, our sensitivity analysis quantifies the total-effect index (S_T_=0.72), followed by edit-rate tolerance (S_T_=0.34), though confidence intervals overlap. The scenario analysis finding that reducing governance from 8-28 weeks to 4 weeks produced d_z_=2.51 suggests governance streamlining may be among the more impactful interventions within the tested set, but the effect should be interpreted in light of the aggressive intervention magnitude (Table S8).

**Abandonment dynamics.** Edit-rate tolerance showed the largest total-effect index for abandonment rate (S_T_=0.88), with abandonment friction threshold a secondary contributor (S_T_=0.28), though uncertainty remains substantial. Empirical studies document that for some users, editing AI-generated notes takes longer than typing, causing net utility to collapse (main Reference 13), and that editing burden is a primary driver of discontinuation. Our model formalizes this as a cumulative friction mechanism where users who exceed individual tolerance thresholds abandon permanently.

**Burnout and champion dynamics.** Burnout reduction calibration draws on Olson and colleagues’ multi-site study documenting a substantial burnout decrease with ambient documentation (main Reference 17), but the High Burnout Crisis scenario showed a near-zero adoption effect. The Champion Program yielded only a modest gain, and the optimistic champion multiplier plus absence of detractors suggest peer-program effects are exploratory rather than definitive. The 2^3^ factorial analysis identified a significant governance x integration interaction (Table S19), while the three-way interaction and other two-way interactions were not significant. Confirming real-world interaction effects would require factorial empirical studies that directly estimate interaction terms.

**Safety implications.** The model’s edit-rate tolerance and abandonment friction parameters operationalize the workload associated with editing AI-generated notes. That workload is also a safety gate: increased editing burden reflects the effort required to detect and correct errors or hallucinations. The model does not distinguish friction from careful review versus friction from poor integration, and both contribute to abandonment in the same way despite different safety implications. Implementation studies could pair integration metrics with safety assurance measures such as accuracy monitoring, feedback loops, and review processes (main Reference 21). The AMA’s finding that 47% of physicians cite increased FDA oversight as the top action needed to increase AI confidence (main Reference 16) indicates that regulatory clarity may be as important as speed.

#### S10.5 Extended Future Directions

Multi-site comparative studies could estimate parameters such as governance delay and edit-rate tolerance that our sensitivity analysis identified as influential but poorly characterized. Comparative evaluations with graded governance streamlining could test dose-response effects implied by the aggressive fast-track scenario. Extending the model to incorporate feedback between adoption levels and technology improvement, with vendors investing in integration quality as market penetration increases, could capture co-evolutionary dynamics absent from the current specification. The model could incorporate learning-curve dynamics documented by Moura and colleagues, which show an initial adjustment period followed by improvements in documentation workload in hybrid ambient programs (main Reference 14). The framework could support prospective exploration of implementation strategies before deployment, allowing health systems to prioritize empirical evaluations of high-impact candidates.

### S11. Extended Methods

#### S11.1 Edit-Rate Tolerance

Edit-rate tolerance represents the maximum fraction of AI-generated documentation requiring physician correction before a clinician considers the tool unacceptable. Each clinician’s tolerance is drawn from Normal(mean=0.45, SD=0.08), consistent with qualitative findings that users abandon when approximately 40% or more of notes require substantive editing (main Reference 13). During trials, exceeding individual tolerance triggers abandonment; post-adoption, excess edit rates contribute to a friction severity score that probabilistically drives reconsideration.

#### S11.2 Burnout Dynamics

Each clinician is initialized with a burnout score drawn from Beta(3.0, 2.0) (mean ~0.6, reflecting the 62.8% physician burnout prevalence reported in AMA surveys; main Reference 16). Burnout influences adoption through psychotype-specific sensitivity multipliers: high-need early adopters have elevated burnout sensitivity (1.5x), while resistant non-adopters have minimal sensitivity (0.3x). Active use reduces burnout by 0.005 per encounter, calibrated so that ~10 weekly encounters over 30 days produces approximately the 13-percentage-point burnout reduction observed by Olson and colleagues (main Reference 17). The High Burnout Crisis scenario shifts baseline burnout to Normal(0.70, 0.10).

#### S11.3 Habit Formation

Active users develop usage habits that provide probabilistic protection against abandonment, with habit strength growing weekly (rate=0.08), a short early-trial window before protection (8 weeks), and a cap at 0.70.

#### S11.4 Organizational Outcome Classification

At simulation end, organizations are classified based on deployment phase: *success* (full_deployment), *scaling* (actively expanding beyond pilot), *pilot stagnation* (pilot phase with retention 0.20-0.55, unable to meet the 0.55 scale threshold), *stalled* (governance or security phases not yet resolved), *failure* (terminated after pilot retention <0.20), and *unknown* (other states). Pilot stagnation thresholds are model-defined: retention floor 0.20 for immediate termination, ceiling 0.55 for immediate scaling, weekly exit hazard 0.02 otherwise, with 0.30 probability of scaling on exit.

#### S11.5 Pilot Stagnation Dynamics

Operationally, pilot stagnation denotes retention between 0.20 and 0.55, with immediate exit if retention rises above 0.55 (scale) or falls below 0.20 (terminate); otherwise a weekly exit hazard of 0.02 applies with a 0.30 probability of scaling on exit. Pilot stagnation is a latent model state inferred by thresholds rather than an observed organizational classification. The comparison to industry reports that many AI projects do not progress beyond pilot is directional rather than definitive; definitions of stalled pilots vary across sources, and the thresholds here are model-defined.

#### S11.6 Implementation

The model was implemented in Python using NumPy for numerical operations, NetworkX for network construction, and Pydantic for configuration validation. Reported analyses were executed in Python 3.12 on macOS. All simulations used explicit random seeding for reproducibility. Analysis code, configuration files, and model documentation will be deposited in a public GitHub repository upon acceptance.

### **References** (Supplementary)

*Note: Numbered references below are specific to supplementary methods. In-text citations using “main Reference X” refer to the main manuscript reference list.*

1. Barnett ML, Landon BE, O’Malley AJ, Keating NL, Christakis NA. Mapping physician networks with self-reported and administrative data. Health Serv Res. 2011;46(5):1592-1609. doi:10.1111/j.1475-6773.2011.01262.x
2. Borracci RA, Giorgi MA. Agent-based model of diffusion of medical innovations. Int J Med Inform. 2018;114:80-87. doi:10.1016/j.ijmedinf.2018.03.015
3. Donohue JM, Guclu H, Gellad WF, et al. Influence of peer networks on physician adoption of new drugs. PLoS One. 2018;13(10):e0204826. doi:10.1371/journal.pone.0204826
4. Greenhalgh T, Robert G, Macfarlane F, Bate P, Kyriakidou O. Diffusion of innovations in service organizations: systematic review and recommendations. Milbank Q. 2004;82(4):581-629. doi:10.1111/j.0887-378X.2004.00325.x
5. Grimm V, Railsback SF, Vincenot CE, et al. The ODD protocol for describing agent-based and other simulation models: a second update to improve clarity, replication, and structural realism. J Artif Soc Soc Simul. 2020;23(2):7. doi:10.18564/jasss.4259
6. Haberle TH, Cleveland C, Snow GL, et al. The impact of Nuance DAX ambient listening AI documentation: a cohort study. J Am Med Inform Assoc. 2024;31(4):975-979. doi:10.1093/jamia/ocae022
7. Olson KD, Meeker D, Troup M, et al. Use of ambient AI scribes to reduce administrative burden and professional burnout among clinicians. JAMA Netw Open. 2025;8(10):e2534976. doi:10.1001/jamanetworkopen.2025.34976
8. Rogers EM. Diffusion of Innovations. 5th ed. New York, NY: Free Press; 2003.
9. Shah SJ, Crowell T, Jeong Y, et al. Physician perspectives on ambient AI scribes: a qualitative study. JAMA Netw Open. 2025;8(3):e251904. doi:10.1001/jamanetworkopen.2025.1904
10. Sobol’ IM. Global sensitivity indices for nonlinear mathematical models and their Monte Carlo estimates. Math Comput Simul. 2001;55(1-3):271-280. doi:10.1016/S0378-4754(00)00270-6
