## Supplementary Results for "An Agent-Based Modeling Framework for Healthcare AI Adoption: Application to Ambient Clinical Documentation"

### S1. Complete Sensitivity Analysis Results

#### Table S9. Sobol Sensitivity Indices for Final Adoption Rate

| Parameter | S_1_ | S_1_ 95% CI | S_T_ | S_T_ 95% CI |
| --- | --- | --- | --- | --- |
| governance_delay_mean | 0.567 | ±0.264 | **0.716** | ±0.272 |
| edit_rate_tolerance_mean | 0.264 | ±0.168 | **0.337** | ±0.114 |
| burnout_reduction_per_use | 0.092 | ±0.127 | 0.151 | ±0.068 |
| peer_influence_coefficient | -0.007 | ±0.102 | 0.143 | ±0.066 |
| innovation_propensity_alpha | -0.033 | ±0.126 | 0.128 | ±0.050 |
| abandonment_friction_threshold | 0.126 | ±0.110 | 0.123 | ±0.042 |
| champion_influence_multiplier | -0.024 | ±0.129 | 0.120 | ±0.043 |
| scale_threshold | -0.045 | ±0.127 | 0.117 | ±0.048 |

*Note: Negative S_1_ values reflect sampling noise at N=64 base samples. Indices should be interpreted as screening-level and ordinal; bold indicates S_T_ > 0.30.*

#### Table S10. Sobol Sensitivity Indices for Abandonment Rate

| Parameter | S_1_ | S_1_ 95% CI | S_T_ | S_T_ 95% CI |
| --- | --- | --- | --- | --- |
| edit_rate_tolerance_mean | 0.748 | ±0.351 | **0.877** | ±0.285 |
| burnout_reduction_per_use | 0.120 | ±0.189 | **0.384** | ±0.184 |
| peer_influence_coefficient | -0.052 | ±0.198 | **0.327** | ±0.137 |
| innovation_propensity_alpha | -0.030 | ±0.198 | **0.322** | ±0.136 |
| governance_delay_mean | -0.012 | ±0.211 | **0.305** | ±0.133 |
| abandonment_friction_threshold | 0.196 | ±0.165 | 0.283 | ±0.104 |
| scale_threshold | 0.019 | ±0.176 | 0.280 | ±0.132 |
| champion_influence_multiplier | 0.023 | ±0.168 | 0.264 | ±0.115 |

*Note: edit_rate_tolerance_mean shows the largest total-effect index on abandonment rate; indices should be interpreted as screening-level with wide confidence intervals.*

#### Sensitivity Analysis Configuration

- **Sampling method**: Saltelli extension of Sobol sequences
- **Base samples (N)**: 64
- **Total parameter sets**: 1,152 [N × (2k + 2) where k=8 parameters; 2,304 simulations with two replicates each]
- **Scale**: 50,000 clinicians
- **Simulation horizon**: 104 weeks
- **Bootstrap resamples for CI**: 100 (SALib default)

*Note: Champion-rate Sobol indices were non-estimable in this run (near-zero variance) and are omitted from the reported tables.*

**Network topology note:** The sensitivity analysis varied eight continuous parameters but did not vary network topology (type, k, or rewiring probability). Because peer influence is a key adoption driver and network structure mediates peer effects, adoption dynamics may differ under alternative topologies (e.g., scale-free or clustered networks). The model supports multiple network types (small_world, random, clustered), but systematic topology sensitivity analysis was not conducted and remains future work.

### S2. Pattern-Oriented Model Plausibility Results

*Note: Plausibility check results (****Tables S11-S13****) are from a separate run set using different random seeds than the emergent analysis (****Tables S15-S18****). Inflection timing therefore differs across run sets; plausibility runs yielded a mean inflection at week 91.3.*

#### Table S11. Complete Plausibility Check Results

| Tier | Pattern | Expected | Observed | Criterion | Status |
| --- | --- | --- | --- | --- | --- |
| **MICRO** | Psychotype ordering | EA → P → LA → NA | EA → P → LA → NA | Spearman ρ ≥ 0.9 | **PASS** (ρ=1.0) |
| **MESO** | Pilot-to-scale attrition | 35-45% | 35.5% | Within range | **PASS** |
| **MACRO** | User distribution | See below | See below | Each category in range | **PASS** |
| **MACRO** | Specialty ordering | PC > SM > Surg > EM | PC > SM > Surg > EM | Spearman ρ ≥ 0.8 | **PASS** (ρ=1.0) |
| **MACRO** | Adoption curve inflection | Week 52-104 | Week 91.3 | Within range | **PASS** |
| **MACRO** | Mature user dropout | 5-15% | 7.7% | Within range | **PASS** |

**Overall**: 6/6 tests passed (100%)

#### Table S12. User Distribution Plausibility Detail

| Category | Expected Range | Target | Observed | Status |
| --- | --- | --- | --- | --- |
| High users (>78% encounters) | 35-50% | 40% | 35.2% | PASS |
| Medium users (30-78%) | 20-35% | 25% | 29.5% | PASS |
| Low users (<30%) | 15-30% | 25% | 23.1% | PASS |
| Abandoned | 5-15% | 9% | 12.1% | PASS |

*Note: Observed distributions fall within acceptance bands for the Stanford qualitative targets, with modest deviations from target values.*

#### Table S13. Psychotype Adoption Timing (MICRO Plausibility Check)

| Psychotype | Mean Week of First Trial | Expected Order |
| --- | --- | --- |
| High-Need Early Adopter | 61.5 | 1 (earliest) |
| Evidence-Driven Pragmatist | 63.1 | 2 |
| Compliance-Oriented Late Adopter | 83.8 | 3 |
| Resistant Non-Adopter | 84.1 | 4 (latest) |

*Spearman ρ = 1.0 (perfect ordering preserved)*

#### Plausibility Check Interpretation

The model reproduces: - **Qualitative adoption dynamics**: S-curve trajectories with appropriate inflection timing - **Psychotype ordering**: Innovation propensity translates correctly to adoption timing - **Usage intensity distribution**: Within empirical acceptance bands - **Specialty ordering**: Correct ordering with ρ=1.0 - **Pilot attrition**: 35.5%, within the 35-45% acceptance band - **Mature user dropout**: Within empirical benchmark range (5-15%)

These results indicate that the calibrated model aligns directionally with prespecified pattern-oriented criteria while retaining the intended qualitative dynamics.

### S3. Detailed Scenario Comparison Results

#### Table S14. Full Scenario Comparison Statistics

| Scenario | n | Baseline Mean | Intervention Mean | Δ (95% CI) | d_z_ |
| --- | --- | --- | --- | --- | --- |
| Fast-Track Governance | 30 | 0.376 | 0.514 | +0.139 [0.119, 0.158] | 2.51 |
| Champion Program | 30 | 0.376 | 0.386 | +0.010 [0.004, 0.017] | 0.55 |
| Better EHR Integration | 30 | 0.376 | 0.394 | +0.018 [0.006, 0.032] | 0.55 |
| Reduced Resistance (proxy) | 30 | 0.376 | 0.416 | +0.040 [0.034, 0.048] | 2.08 |
| High Burnout Crisis | 30 | 0.376 | 0.380 | +0.005 [-0.013, 0.021] | 0.09 |
| Combined Intervention | 30 | 0.376 | 0.575 | +0.200 [0.187, 0.213] | 5.23 |

*Δ values are paired mean differences with 95% bootstrap confidence intervals (1,000 resamples). P-values from paired t-tests were computed but are de-emphasized to avoid overinterpreting stochastic simulation significance.*

#### Interaction Effects

| Individual Effects Sum | Combined Effect | Interaction Ratio |
| --- | --- | --- |
| +16.7pp (Fast Gov + Champion + Better Integration) | +20.0pp | **1.20** |

The combined intervention exceeds the sum of individual effects by 20% (3.3pp). A full 2^3^ factorial analysis (Supplementary Methods S9, Table S19, **Figure S4**) provides formal interaction tests for these effects.

### S4. Emergent Analysis Detailed Results

*Note: Emergent analysis results (****Tables S15-S18****) are from the primary analysis run set (n=30 realizations, 50,000 clinicians). These results differ from validation runs (Section S2) due to different random seeds.*

#### Table S15. Inflection Point Statistics (n=30 realizations)

| Statistic | Value |
| --- | --- |
| Mean inflection week | 51.0 |
| SD | 28.6 |
| Median | 43.9 |
| Min | 19.0 |
| Max | 90.3 |
| Inflection points per trajectory | 3.0 (mean) |

#### Table S16. Organizational Outcome Distribution

| Outcome | Definition | Proportion | 95% CI |
| --- | --- | --- | --- |
| Pilot Stagnation | Stuck in pilot/early scaling | 48.9% | [48.2%, 49.8%] |
| Failure | Terminated deployment | 3.4% | [2.5%, 4.6%] |
| Success | Full deployment achieved | 28.2% | [27.1%, 29.3%] |
| Stalled | Partial adoption plateau | 6.9% | [6.6%, 7.4%] |
| Scaling | Active expansion | 9.4% | [8.9%, 9.9%] |
| Unknown | Indeterminate | 3.1% | [2.7%, 3.1%] |

#### Table S17. Champion and Detractor Emergence

| Metric | Mean | SD | Min | Max |
| --- | --- | --- | --- | --- |
| Champions (n) | 3,638 | 948 | 1,130 | 4,907 |
| Champions (%) | 7.3% | 1.9% | 2.3% | 9.8% |
| Detractors (n) | 0 | 0 | 0 | 0 |
| Detractors (%) | 0.0% | 0.0% | 0.0% | 0.0% |

*Note: The absence of detractors is a model limitation. The current model structure routes dissatisfied users to abandonment rather than representing persistent vocal critics who remain in the system and actively discourage peers. Real-world deployments likely include such detractors, whose influence on adoption dynamics warrants future modeling attention.*

#### Table S18. Specialty-Specific Adoption Rates (n=30 realizations)

| Specialty | Mean Adoption | SD | Abandonment Rate | Champion Rate |
| --- | --- | --- | --- | --- |
| Primary Care | 39.8% | 3.0% | 3.1% | 10.4% |
| Specialty Medical | 37.9% | 3.2% | 5.0% | 7.1% |
| Surgical | 34.9% | 3.4% | 8.0% | 3.9% |
| Emergency | 31.2% | 3.7% | 11.5% | 2.3% |

*Ordering by adoption rate: Primary Care > Specialty Medical > Surgical > Emergency* *Ordering by abandonment: Emergency > Surgical > Specialty Medical > Primary Care (inverse)*

### S5. Robustness and Convergence

#### Stochastic Stability

To assess stochastic stability, we examined coefficient of variation (CV) across 30 realizations:

| Output Metric | Mean | SD | CV |
| --- | --- | --- | --- |
| Final adoption rate | 0.374 | 0.030 | 0.08 |
| Time to 10% adoption | 60.5 weeks | 5.6 | 0.09 |
| Champion count | 3,638 | 948 | 0.26 |
| Abandonment rate | 0.055 | 0.026 | 0.47 |

CV values of 0.08-0.47 indicate moderate stochastic variability, appropriate for a model capturing path-dependent adoption dynamics where early random events influence subsequent trajectories.

*Note: Values in this table are from the emergent analysis run set (n=30 realizations, independent seeds). The paired scenario baselines (Table S14) report 0.376 due to different seed matching.*

#### Scale Sensitivity

Scale sensitivity was not re-evaluated in the current run, so prior exploratory results are omitted here to avoid mixing generations of outputs.

### S6. Supplementary Figures

*Note: Full-resolution figures are provided as separate files.*

- **Figure S1**: Psychotype validation analysis showing (A) activation timing by adopter type and (B) adoption rates by psychotype, demonstrating alignment with Rogers diffusion theory (Spearman ρ = 1.0)
- **Figure S2**: Organizational outcome distribution showing (A) proportions across six outcome categories and (B) variability across realizations
- **Figure S3**: Parameter sensitivity heatmap showing total-order Sobol indices across output metrics
- **Figure S4**: 2^3^ factorial interaction analysis showing (A) three-factor interaction plot and (B-D) marginal interaction plots for governance, champion, and integration factors (see Supplementary Methods S9, Table S19)

### References (Supplementary Results)

Data sources for validation targets (see main References):

- Intermountain DAX cohort study (main Reference 5): 41% pilot attrition target
- Stanford qualitative study (main Reference 13): Usage distribution targets
- Rogers diffusion categories (main Reference 10): Psychotype ordering
- Kaiser Permanente deployment report (main Reference 2): Adoption trajectory benchmarks
